# Outcomes in Patients with Acute Hypoxemic Respiratory Failure Secondary to COVID-19 Treated with Noninvasive Respiratory Support versus Invasive Mechanical Ventilation

**DOI:** 10.1101/2022.12.19.22283704

**Authors:** Julia M Fisher, Vignesh Subbian, Patrick Essay, Sarah Pungitore, Edward J Bedrick, Jarrod M Mosier

## Abstract

**Purpose:** The goal of this study was to compare noninvasive respiratory support to invasive mechanical ventilation as the initial respiratory support in COVID-19 patients with acute hypoxemic respiratory failure.

**Methods:** All patients admitted to a large healthcare network with acute hypoxemic respiratory failure associated with COVID-19 and requiring respiratory support were eligible for inclusion. We compared patients treated initially with noninvasive respiratory support (noninvasive positive pressure ventilation by facemask or high flow nasal oxygen) with patients treated initially with invasive mechanical ventilation. The primary outcome was time-to-in-hospital death analyzed using an inverse probability of treatment weighted Cox model adjusted for potential confounders. Secondary outcomes included unweighted and weighted assessments of mortality, lengths-of-stay (intensive care unit and hospital) and time-to-intubation.

**Results:** Over the study period, 2354 patients met inclusion criteria. Nearly half (47%) received invasive mechanical ventilation first and 53% received initial noninvasive respiratory support. There was an overall 38% in-hospital mortality (37% for invasive mechanical ventilation and 39% for noninvasive respiratory support). Initial noninvasive respiratory support was associated with an increased hazard of death compared to initial invasive mechanical ventilation (HR: 1.61, p < 0.0001, 95% CI: 1.33 - 1.94). However, patients on initial noninvasive respiratory support also experienced an increased hazard of leaving the hospital sooner, but the hazard ratio waned with time (HR: 0.97, p < 0.0001, 95% CI: 0.96 - 0.98).

**Conclusion:** These data show that the COVID-19 patients with acute hypoxemic respiratory failure initially treated with noninvasive respiratory support had an increased hazard of in-hospital death.

## Introduction

The optimal management strategy for patients with acute hypoxemic respiratory failure due to SARS-CoV-2 infection has undergone particular interest and scrutiny. Early, intense discussion occurred in the published literature and on social media. High failure rates with noninvasive positive pressure ventilation (NIPPV) during the SARS-CoV outbreak in 2003,^1^ concerns over aerosol transmission with noninvasive support strategies,^2–8^ potential novelty of respiratory physiology with COVID-19,^9–18^ impending ventilator shortages,^19–21^ and the high mortality initially reported with invasive mechanical ventilation (IMV)^22, 23^ all factored in the discussion over timing of intubation and the utility of noninvasive strategies.

The current literature on noninvasive respiratory support (NIRS) — NIPPV or high flow nasal oxygen (HFNO) — for patients with COVID-19 report disparate outcomes. Importantly, these studies compare noninvasive strategies to each other, or to conventional oxygen therapy, but not to invasive mechanical ventilation. The goal of this study was to explore the optimal initial treatment for COVID-19 patients admitted with acute hypoxemic respiratory failure by comparing invasive versus noninvasive strategies.

## Methods

### Study Design and Setting

This study is a retrospective cohort observational study using clinical data. Clinical data were obtained from the Banner Health Network clinical data warehouse, covering 26 hospitals across six states in the western United States. Data were extracted for all adult patients (≥18 years) admitted for respiratory failure associated with COVID-19 between January 1, 2020 and January 7, 2021. Data consist of de-identified structured data generated from the Cerner (Cerner Corporation, North Kansas City, MO, USA) electronic health record. This work adheres to the STROBE reporting guidelines and was approved by the University of Arizona (IRB #1907780973) and Banner Health Institutional Review Boards (IRB #483-20-0018).

### Study Participants and Treatment Assignment

We generated seven cohorts using a phenotyping algorithm^24^ based on the sequence of therapies received: (1) IMV only, (2) NIPPV only, (3) HFNO only, (4) NIPPV requiring subsequent IMV, (5) HFNO requiring subsequent IMV, (6) IMV extubated to NIPPV, (7) IMV extubated to HFNO, and (8) evidence of all three treatments but unclear treatment ordering. Those treated with any noninvasive respiratory support modality first were compared to those treated with invasive mechanical ventilation first. Subsequent analyses separated NIPPV and HFNO and included pairwise comparisons of initial IMV, NIPPV, and HFNO.

We estimated the propensity to be given each modality using generalized boosted models and used inverse probability of treatment weighting in the models to account for non-random treatment assignment.^25^ The variables for propensity score estimation included age, body mass index, sex, ethnicity (non-Hispanic, Hispanic), race (white, non-white), respiratory rate and S_P_O_2_/F_I_O_2_ ratio most immediately prior to first treatment, hospital size by American Hospital Association category (small [< 100 beds], medium [100-499 beds], large [> 500 beds]), and either hours from hospital admission to first treatment or hours from ICU admission to first treatment, transformed via the Box-Cox method with negatives.^26^ These variables were additionally included in Cox models to further improve balance between treatment groups.

### Outcomes and Data Analysis

The primary outcome is time-to-in-hospital death. We fit a Cox model from first treatment initiation to death with censoring occurring at hospital discharge. However, there are multiple potential pathways for a patient (1. death, 2. discharge alive [from hospital or ICU], 3. remain in the hospital, and 4. intubation [if on NIRS]). We modeled discharge alive and intubation with three additional hazard models. First, we fit a cause-specific hazard model for time-to-*hospital* discharge alive with death as a competing risk where time zero was set as time of first treatment initiation. For this outcome, we performed a planned sensitivity analysis with time zero set as time of hospital entrance. Second, we fit a cause-specific hazard model for time-to-*ICU* discharge alive (from ICU admission) with death as a competing risk. Only visits with an ICU stay were included in this analysis; thus, results are conditional on having been admitted to the ICU and by nature excluded ICU-level of care patients who remained outside of the ICU. Third, we conducted a competing risks analysis for time-to-*intubation* with death as a competing risk and censoring occurring at hospital discharge. Only visits with known time to event (96.3% of possible visits) and a noninvasive initial treatment were included. The competing risks analysis used a modification of Gray’s test that incorporates inverse probability of treatment weighting.^27^ Statistical significance was judged via the median p-value approach.^28^ Planned sensitivity analyses were performed to assess preprocessing and analysis decisions made for the competing risks (see online supplement for details).

We analyzed unweighted outcomes of mortality, intubation rate, duration of mechanical ventilation in patients who failed initial NIRS treatment, length-of-hospital-stay, and ICU-free days using Fisher’s Exact and Kruskal-Wallis rank sum tests where appropriate. For all outcomes, the predictor of interest was first treatment (noninvasive respiratory support or invasive mechanical ventilation). We assessed if the proportional hazard assumption was violated in the Cox models by including an interaction of time by first treatment. If the interaction was statistically significant at α = 0.05, we report the interaction model; otherwise, we report the proportional hazards model.

Data preprocessing is described in detail in the online supplement. As a retrospective study based on routinely collected EHR data, it is expected that the extent of observed data will be different for different clinical variables.^29–31^ Missing data were handled by using multiple imputation by chained equations (MICE).^32, 33^ For each non-competing risks analysis, we created 50 imputed data sets using all variables in the propensity score construction, the Nelson-Aalen estimate of the available time-to-event data, the time-to-event itself, and event information (e.g., death, hospital discharge alive, ICU discharge alive). Age, body mass index, SpO_2_/FiO_2_, the transformed time-from-hospital admission (or ICU admission) to first treatment, respiratory rate, time-to-event, and the Nelson-Aalen estimate of the time-to-event were imputed via predictive mean matching. Sex, ethnicity, race, the outcome event, hospital size, and first treatment were imputed with logistic regression (two-category variables) or multinomial log-linear models via neural networks (> two category variables) as appropriate. Raw time-to-event was imputed but not used to predict other variables in the MICE algorithm. Instead, for the Cox models and some competing risk models, temporal information was used in the prediction of other missing values via the Nelson-Aalen estimate. For the competing risk models using the median p-value inference approach, no outcome information was allowed to predict other variables in the MICE algorithm. Since each imputed data set had different values for variables used in the propensity score algorithm, we estimated propensity scores for each imputed data set separately. For the Cox models, the propensity scores from a specific imputed data set were used to do inverse probability of treatment weighting with the analysis of that data set,^34^ and results were combined using Rubin’s Rules. For the competing risks analyses, the propensity scores from each imputed data set were handled differently depending on the inference approach. See the supplementary information for further details. All data preprocessing and statistical analyses were done using R version 4.0.4^35^ and included the following packages: twang^34^, survival^36, 37^, survminer,^38^ MICE^32^, xtable,^39^ and tidyverse.^40^

## Results

During the study period, there were 2354 COVID-19 patients that met criteria for inclusion. There were 326 that were not classified by the phenotyping algorithm but received all three treatments. Of the 2028 patients that were reliably classified, invasive mechanical ventilation was used as the first therapy in 947 (47%) patients and noninvasive respiratory support in 1081 (53%), **Figure 1**, **Table 1**. Of those on noninvasive respiratory support, 811 (75%) received NIPPV first and 270 (25%) received HFNO first. There was imbalance in NIRS use among hospitals of different sizes with larger hospitals disproportionately using NIPPV first (large hospitals 81% [433/536], medium hospitals 70% [346/495], small hospitals 64% [32/50]) **Table 1**. For the 326 unclassifiable records, the first treatment was imputed using multiple imputation. Since treatment assignments may have varied between imputed data sets, demographics for this group are reported separately in the online supplement.

**Figure 1:**
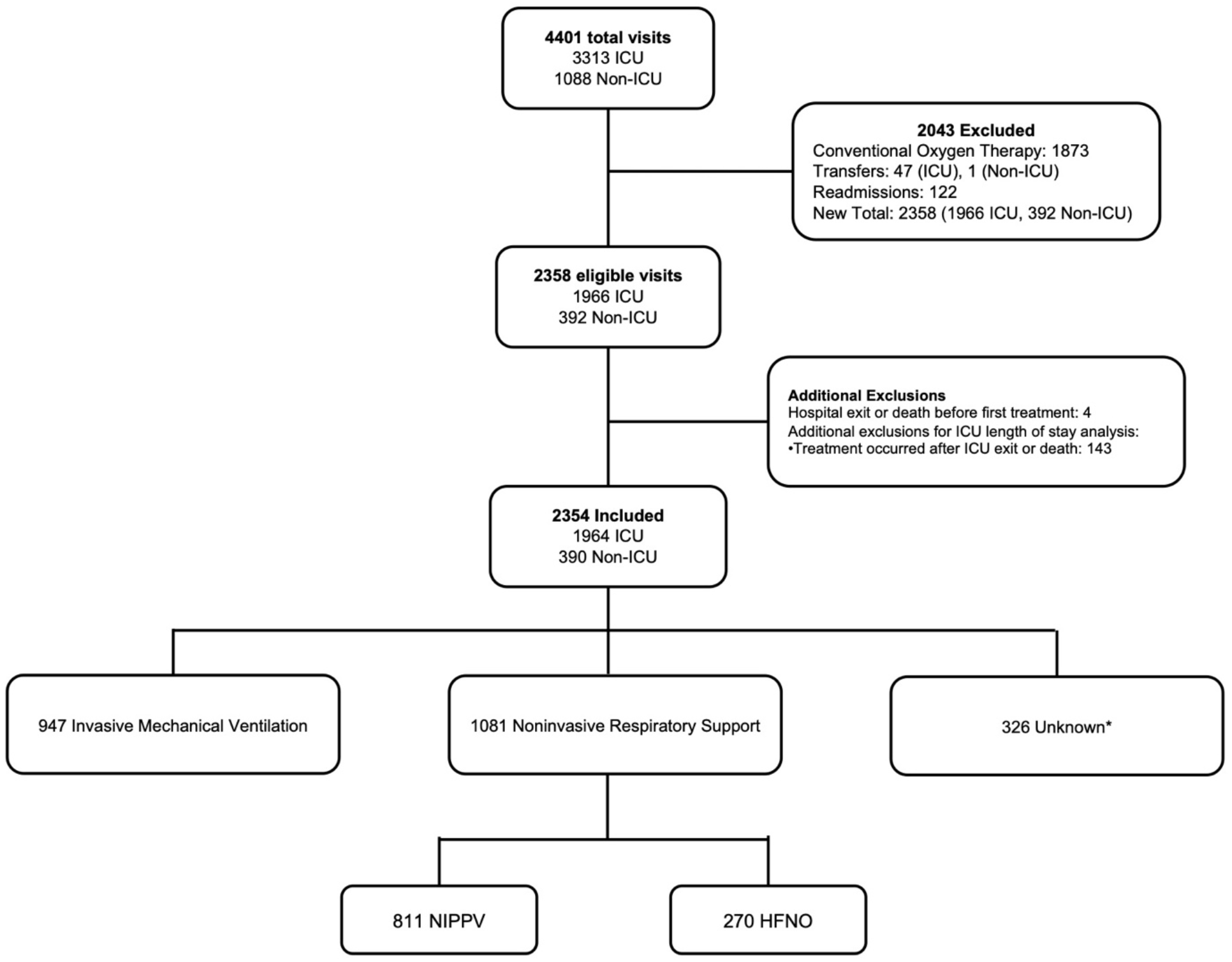
STROBE Statement 326 participants had evidence of high-flow nasal oxygen, noninvasive positive pressure ventilation, and invasive mechanical ventilation but were unable to be reliably classified by the phenotyping algorithm because of nonsensical time stamps for timing of therapies. See text and online supplement for further description and demographics of this group. “Additional exclusions” includes 4 participants that were excluded because of nonsensical hospital discharge time stamps, and an additional group of patients excluded only from the model assessing time-to-ICU exit.

**Table 1:**
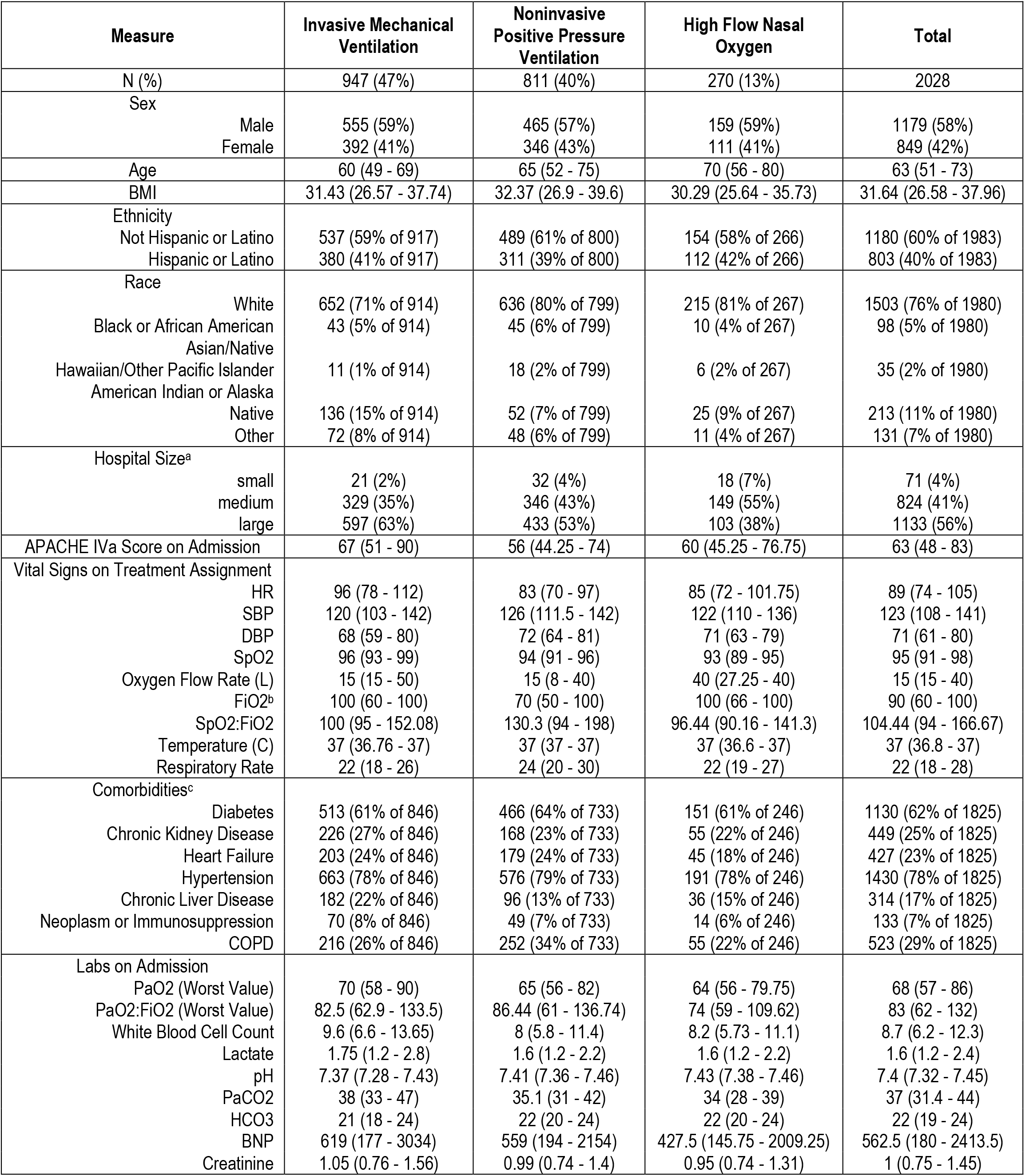
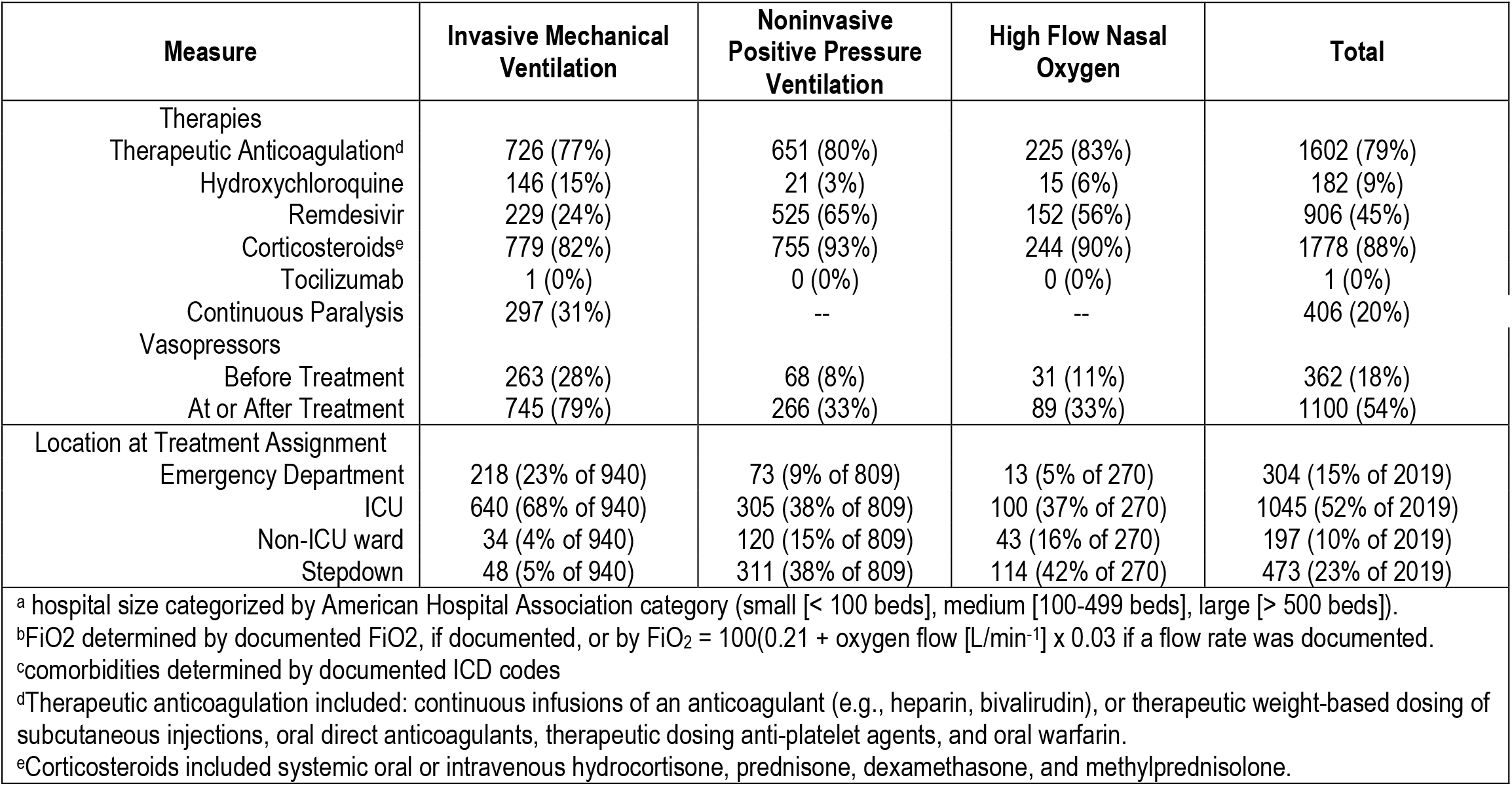
Demographics

| Measure | Invasive Mechanical Ventilation | Noninvasive Positive Pressure Ventilation | High Flow Nasal Oxygen | Total |
| --- | --- | --- | --- | --- |
| N (%) | 947 (47%) | 811 (40%) | 270 (13%) | 2028 |
| Sex |  |  |  |  |
| Male | 555 (59%) | 465 (57%) | 159 (59%) | 1179 (58%) |
| Female | 392 (41%) | 346 (43%) | 111 (41%) | 849 (42%) |
| Age | 60 (49 - 69) | 65 (52 - 75) | 70 (56 - 80) | 63 (51 - 73) |
| BMI | 31.43 (26.57 - 37.74) | 32.37 (26.9 - 39.6) | 30.29 (25.64 - 35.73) | 31.64 (26.58 - 37.96) |
| Ethnicity |  |  |  |  |
| Not Hispanic or Latino | 537 (59% of 917) | 489 (61% of 800) | 154 (58% of 266) | 1180 (60% of 1983) |
| Hispanic or Latino | 380 (41% of 917) | 311 (39% of 800) | 112 (42% of 266) | 803 (40% of 1983) |
| Race |  |  |  |  |
| White | 652 (71% of 914) | 636 (80% of 799) | 215 (81% of 267) | 1503 (76% of 1980) |
| Black or African American | 43 (5% of 914) | 45 (6% of 799) | 10 (4% of 267) | 98 (5% of 1980) |
| Asian/Native Hawaiian/Other Pacific Islander | 11 (1% of 914) | 18 (2% of 799) | 6 (2% of 267) | 35 (2% of 1980) |
| American Indian or Alaska Native | 136 (15% of 914) | 52 (7% of 799) | 25 (9% of 267) | 213 (11% of 1980) |
| Other | 72 (8% of 914) | 48 (6% of 799) | 11 (4% of 267) | 131 (7% of 1980) |
| Hospital Size<sup>a</sup> |  |  |  |  |
| small | 21 (2%) | 32 (4%) | 18 (7%) | 71 (4%) |
| medium | 329 (35%) | 346 (43%) | 149 (55%) | 824 (41%) |
| large | 597 (63%) | 433 (53%) | 103 (38%) | 1133 (56%) |
| APACHE IVa Score on Admission | 67 (51 - 90) | 56 (44.25 - 74) | 60 (45.25 - 76.75) | 63 (48 - 83) |
| Vital Signs on Treatment Assignment |  |  |  |  |
| HR | 96 (78 - 112) | 83 (70 - 97) | 85 (72 - 101.75) | 89 (74 - 105) |
| SBP | 120 (103 - 142) | 126 (111.5 - 142) | 122 (110 - 136) | 123 (108 - 141) |
| DBP | 68 (59 - 80) | 72 (64 - 81) | 71 (63 - 79) | 71 (61 - 80) |
| SpO2 | 96 (93 - 99) | 94 (91 - 96) | 93 (89 - 95) | 95 (91 - 98) |
| Oxygen Flow Rate (L) | 15 (15 - 50) | 15 (8 - 40) | 40 (27.25 - 40) | 15 (15 - 40) |
| FiO2<sup>b</sup> | 100 (60 - 100) | 70 (50 - 100) | 100 (66 - 100) | 90 (60 - 100) |
| SpO2:FiO2 | 100 (95 - 152.08) | 130.3 (94 - 198) | 96.44 (90.16 - 141.3) | 104.44 (94 - 166.67) |
| Temperature (C) | 37 (36.76 - 37) | 37 (37 - 37) | 37 (36.6 - 37) | 37 (36.8 - 37) |
| Respiratory Rate | 22 (18 - 26) | 24 (20 - 30) | 22 (19 - 27) | 22 (18 - 28) |
| Comorbidities<sup>c</sup> |  |  |  |  |
| Diabetes | 513 (61% of 846) | 466 (64% of 733) | 151 (61% of 246) | 1130 (62% of 1825) |
| Chronic Kidney Disease | 226 (27% of 846) | 168 (23% of 733) | 55 (22% of 246) | 449 (25% of 1825) |
| Heart Failure | 203 (24% of 846) | 179 (24% of 733) | 45 (18% of 246) | 427 (23% of 1825) |
| Hypertension | 663 (78% of 846) | 576 (79% of 733) | 191 (78% of 246) | 1430 (78% of 1825) |
| Chronic Liver Disease | 182 (22% of 846) | 96 (13% of 733) | 36 (15% of 246) | 314 (17% of 1825) |
| Neoplasm or Immunosuppression | 70 (8% of 846) | 49 (7% of 733) | 14 (6% of 246) | 133 (7% of 1825) |
| COPD | 216 (26% of 846) | 252 (34% of 733) | 55 (22% of 246) | 523 (29% of 1825) |
| Labs on Admission |  |  |  |  |
| PaO2 (Worst Value) | 70 (58 - 90) | 65 (56 - 82) | 64 (56 - 79.75) | 68 (57 - 86) |
| PaO2:FiO2 (Worst Value) | 82.5 (62.9 - 133.5) | 86.44 (61 - 136.74) | 74 (59 - 109.62) | 83 (62 - 132) |
| White Blood Cell Count | 9.6 (6.6 - 13.65) | 8 (5.8 - 11.4) | 8.2 (5.73 - 11.1) | 8.7 (6.2 - 12.3) |
| Lactate | 1.75 (1.2 - 2.8) | 1.6 (1.2 - 2.2) | 1.6 (1.2 - 2.2) | 1.6 (1.2 - 2.4) |
| pH | 7.37 (7.28 - 7.43) | 7.41 (7.36 - 7.46) | 7.43 (7.38 - 7.46) | 7.4 (7.32 - 7.45) |
| PaCO2 | 38 (33 - 47) | 35.1 (31 - 42) | 34 (28 - 39) | 37 (31.4 - 44) |
| HCO3 | 21 (18 - 24) | 22 (20 - 24) | 22 (20 - 24) | 22 (19 - 24) |
| BNP | 619 (177 - 3034) | 559 (194 - 2154) | 427.5 (145.75 - 2009.25) | 562.5 (180 - 2413.5) |
| Creatinine | 1.05 (0.76 - 1.56) | 0.99 (0.74 - 1.4) | 0.95 (0.74 - 1.31) | 1 (0.75 - 1.45) |
| Therapies |  |  |  |  |
| Therapeutic Anticoagulation<sup>d</sup> | 726 (77%) | 651 (80%) | 225 (83%) | 1602 (79%) |
| Hydroxychloroquine | 146 (15%) | 21 (3%) | 15 (6%) | 182 (9%) |
| Remdesivir | 229 (24%) | 525 (65%) | 152 (56%) | 906 (45%) |
| Corticosteroids<sup>e</sup> | 779 (82%) | 755 (93%) | 244 (90%) | 1778 (88%) |
| Tocilizumab | 1 (0%) | 0 (0%) | 0 (0%) | 1 (0%) |
| Continuous Paralysis | 297 (31%) | -- | -- | 406 (20%) |
| Vasopressors |  |  |  |  |
| Before Treatment | 263 (28%) | 68 (8%) | 31 (11%) | 362 (18%) |
| At or After Treatment | 745 (79%) | 266 (33%) | 89 (33%) | 1100 (54%) |
| Location at Treatment Assignment |  |  |  |  |
| Emergency Department | 218 (23% of 940) | 73 (9% of 809) | 13 (5% of 270) | 304 (15% of 2019) |
| ICU | 640 (68% of 940) | 305 (38% of 809) | 100 (37% of 270) | 1045 (52% of 2019) |
| Non-ICU ward | 34 (4% of 940) | 120 (15% of 809) | 43 (16% of 270) | 197 (10% of 2019) |
| Stepdown | 48 (5% of 940) | 311 (38% of 809) | 114 (42% of 270) | 473 (23% of 2019) |

<sup>a</sup> hospital size categorized by American Hospital Association category (small [ $< 100$ beds], medium [100-499 beds], large [ $> 500$ beds]).
<sup>b</sup>FiO<sub>2</sub> determined by documented FiO<sub>2</sub>, if documented, or by $\text{FiO}_2 = 100(0.21 + \text{oxygen flow [L/min}^{-1}] \times 0.03$ if a flow rate was documented.
<sup>c</sup>comorbidities determined by documented ICD codes
<sup>d</sup>Therapeutic anticoagulation included: continuous infusions of an anticoagulant (e.g., heparin, bivalirudin), or therapeutic weight-based dosing of subcutaneous injections, oral direct anticoagulants, therapeutic dosing anti-platelet agents, and oral warfarin.
<sup>e</sup>Corticosteroids included systemic oral or intravenous hydrocortisone, prednisone, dexamethasone, and methylprednisolone.

All-cause in-hospital mortality was 38%. For those on IMV first, mortality was 37%. For those treated with noninvasive respiratory support first and never required intubation, mortality was 29%, but rose to 60% for those that required intubation. There was also an imbalance in mortality rates between NIPPV and HFNO, **Table 2**. Intubation rates for those treated with HFNO and NIPPV were 32% (87/270) and 33% (268/811), respectively.

**Table 2:** Unmatched outcomes

| Measure | Invasive Mechanical Ventilation | Noninvasive Positive Pressure Ventilation | High Flow Nasal Oxygen | Total | P-Value |
| --- | --- | --- | --- | --- | --- |
| All-cause in-ICU mortality |  |  |  |  |  |
| Total | 253 (28% of 891) | 138 (32% of 426) | 75 (57% of 131) | 466 (32% of 1448) | <0.001 |
| Failure | -- | 99 (41% of 244) | 47 (64% of 74) | 146 (46% of 318) | 0.001 |
| Success | -- | 39 (21% of 182) | 28 (49% of 57) | 67 (28% of 239) | <0.001 |
| All-cause hospital mortality |  |  |  |  |  |
| Total | 348 (37% of 947) | 281 (35% of 811) | 140 (52% of 270) | 769 (38% of 2028) | <0.001 |
| Failure | -- | 145 (54% of 268) | 67 (77% of 87) | 212 (60% of 355) | <0.001 |
| Success | -- | 136 (25% of 543) | 73 (40% of 183) | 209 (29% of 726) | <0.001 |
| Failure (intubation rate) | -- | 268 (33% of 811) | 87 (32% of 270) | 355 (33% of 1081) | 0.001 |
| Days to Intubation | -- | 0.24 (0.13 – 1.18) | 0.14 (0.06 – 0.56) | 0.21 (0.11 – 1.05) | <0.001 |
| Duration of mechanical ventilation (days) | -- | 8.08 (3.6 - 13.98) | 8.36 (1.4 - 17.37) | 8.09 (3.34 - 14.91) | 0.516 |
| ICU length of stay (days) |  |  |  |  |  |
| Total | 8.04 (3.49 - 16.75) | 6.2 (2.25 - 12.7) | 7.15 (2.59 - 13.66) | 7.48 (3 - 15.2) | <0.001 |
| Failure | -- | 10.77 (5.02 – 18.03) | 12.26 (3.5 – 19.36) | 10.81 (4.65 – 18.73) | 0.916 |
| Success | -- | 2.68 (1.19 – 5.5) | 3.99 (2.3 – 7.61) | 2.91 (1.33 – 6.04) | 0.014 |
| ICU free days (among those who were in the ICU) |  |  |  |  |  |
| Total | 5.5 (0.56 – 12.59) | 6.69 (2.03 – 11.96) | 3.59 (0.3 – 9.41) | 5.62 (0.99 – 12.22) | 0.003 |
| Failure | -- | 6.97 (1.69 – 12.71) | 4.72 (0.74 – 11.81) | 6.72 (1.43 – 12.43) | 0.115 |
| Success | -- | 5.6 (2.4 – 10.61) | 2.27 (0.14 – 6.53) | 5.03 (1.67 – 9.91) | <0.001 |
| Hospital length of stay (days) |  |  |  |  |  |
| Total | 17.59 (9.47 – 28.23) | 12.32 (6.97 – 21.06) | 11.75 (7.22 – 18.98) | 14.46 (7.95 – 24.24) | <0.001 |
| Failure | -- | 20.17 (13.79 – 29.27) | 17.61 (11.44 – 27.77) | 19.85 (13.22 – 29.02) | 0.157 |
| Success | -- | 9.8 (5.84 - 15.22) | 9.86 (5.93 - 14.8) | 9.82 (5.85 - 15.03) | 0.655 |

Estimates are n (% of column n with available data) for categorical characteristics and median (interquartile range) for continuous characteristics. Inferences for categorical variables are the result of Fisher Exact Tests, with p-values computed via Monte Carlo simulation when necessary for computational efficiency. Inferences for continuous variables come from Kruskal-Wallis rank sum test of group differences.

### NIRS vs IMV

Initial noninvasive respiratory support was associated with an increased hazard of in-hospital death compared to initial invasive mechanical ventilation (HR: 1.61, p < 0.0001, 95% CI:1.33 - 1.94), and there was no significant interaction of treatment and time with this association (HR: 1.00, p = 0.6038, 95% CI: 0.98 - 1.01) **Figure 2**. However, being on initial noninvasive respiratory support was also associated with an early increased hazard of leaving the hospital alive, but this hazard ratio decreased over time due to an interaction between time and treatment. This eventually resulted in the hazard ratio reversing such that initial noninvasive respiratory support was later associated with a decreased hazard of leaving the hospital alive (HR: 0.97, p < 0.0001, 95% CI: 0.96 - 0.98) **FIGURE 2.** This was consistent with the sensitivity analysis where time zero is hospital admission (online supplement). There was also a similar pattern in the difference between initial NIRS and initial IMV in the time-to-ICU discharge alive: early on, initial noninvasive respiratory support was associated with an increased hazard of discharge alive from the ICU, but this hazard ratio decreased over time because of a statistically significant interaction between treatment and time (HR: 0.98, p = 0.0321, 95% CI: 0.97 - 1.00), resulting in no significant differences between non-invasive and invasive initial support at later time points..

**Figure 2:**
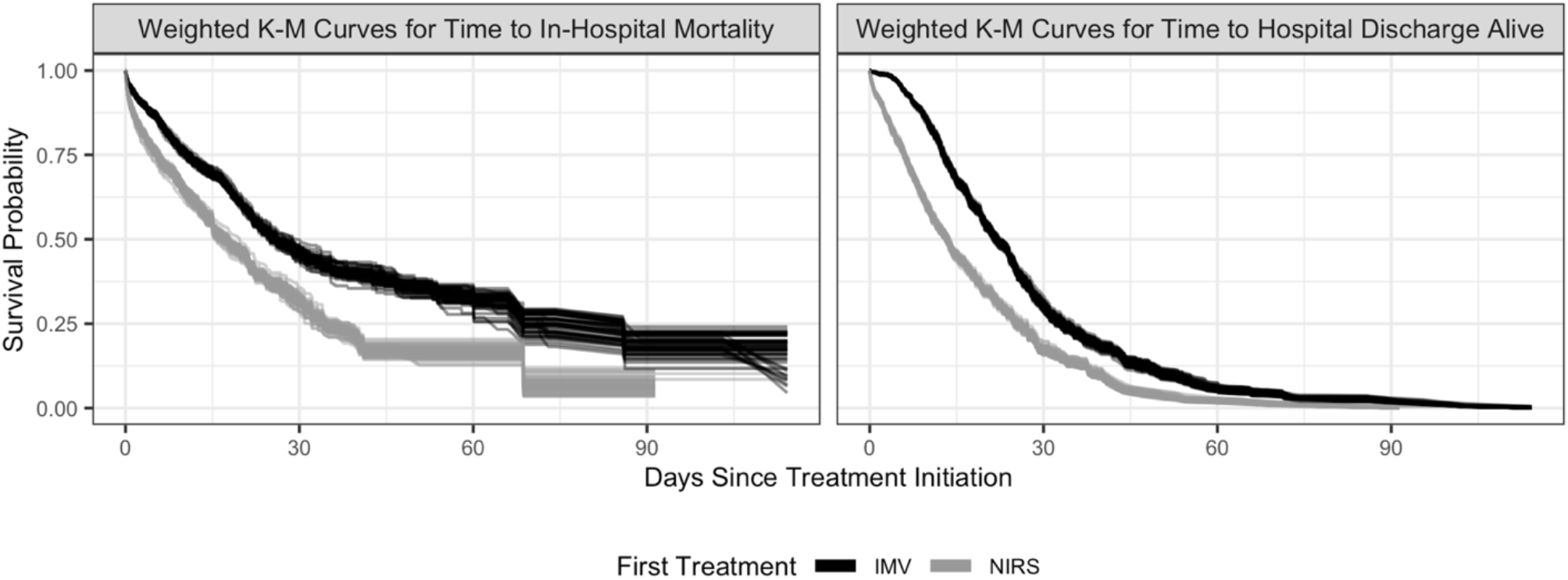
Kaplan-Meier curves for NIRS vs IMV for death (left) and discharge alive (right)

### HFNO vs NIPPV vs IMV

Both HFNO (HR: 2.37, p < 0.0001, 95% CI: 1.85 - 3.03) and NIPPV (HR: 1.44, p = 0.0003, 95% CI: 1.18 - 1.74) were associated with increased hazards of in-hospital mortality compared to invasive mechanical ventilation. HFNO had an increased hazard of in-hospital death compared to NIPPV (HR: 1.65, p = 0.0001, 95% CI: 1.29 - 2.10). The interactions of time and both NIPPV and HFNO were not statistically significant and were excluded from the final model (both p > 0.05).

For time-to-hospital discharge alive, there were significant interactions between treatment and time for both NIRS modalities (time by HFNO HR: 0.96, p = 0.0011, 95% CI: 0.94 - 0.98 and time by NIPPV HR: 0.98, p = 0.0231, 95% CI: 0.97 - 1.00). Similar to the models that grouped NIPPV and HFNO, these models showed increased hazards of discharge alive for both HFNO and NIPPV compared to invasive mechanical ventilation that decreased over time but at different rates (HFNO faster, NIPPV slower), **FIGURE 3.** This resulted in no statistically significant differences between NIPPV and invasive mechanical ventilation at later time points but a decreased hazard of discharge alive for HFNO compared to invasive mechanical ventilation.

**Figure 3:**
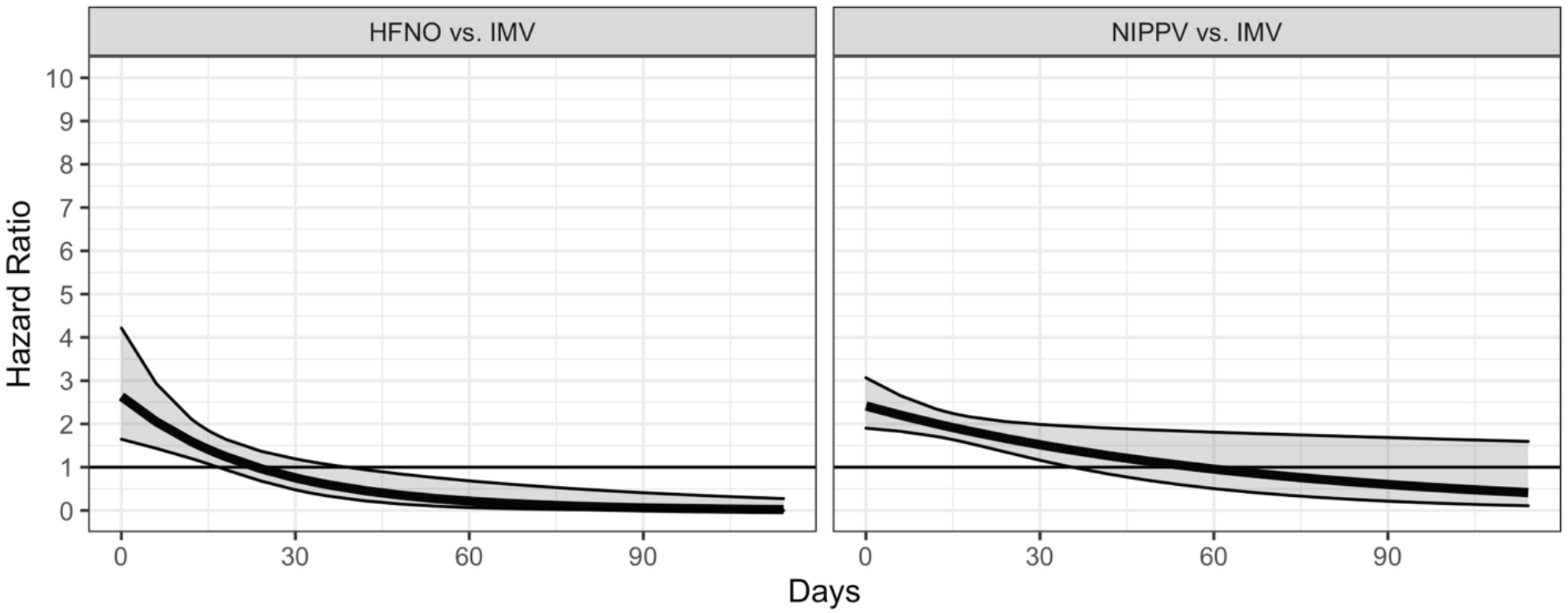
Estimated Hazard Ratios for Time-to-Discharge Alive **HFNO** vs. IMV (left) and **NIPPV** vs. IMV (right) at various points in time. Pointwise 95% confidence intervals included.

Patients who started on HFNO first and were intubated were more likely to be intubated sooner than those who started on NIPPV (p < 0.001). First treatment with both IMV and NIPPV were associated with a higher hazard of ICU discharge alive than first treatment with HFNO (IMV HR: 1.50, p = 0.0226, 95% CI: 1.06 - 2.13; NIPPV HR: 1.84, p = 0.0014, 95% CI: 1.27 - 2.68), and first treatment with NIPPV was associated with a higher hazard of leaving the ICU alive than first treatment with IMV (HR: 1.23, p = 0.0341, 95% CI: 1.02 - 1.48). There was no significant interaction between time and treatment for time-to-ICU discharge alive (time by HFNO HR: 0.99, p = 0.4065, 95% CI: 0.97 - 1.01; time by NIPPV HR: 0.99, p = 0.3047, 95% CI: 0.97 - 1.01), and these interactions were left out of the final model. All competing risk sensitivity analyses showed the same pattern and were also statistically significant (all p < 0.001, see online supplement). In the sensitivity analysis of time-to-hospital discharge alive, the interaction of time with HFNO was statistically significant (p = 0.0055) but the interaction of time with NIPPV was not (p = 0.0848). Despite this, the relationship between therapies with the time by treatment interaction was consistent with the model where time zero was set to first treatment initiation.

## Discussion

Our results show that initial noninvasive respiratory support presents a double-edged sword for COVID-19 patients with acute respiratory failure. Patients supported first by NIRS experience a greater hazard of in-hospital death compared to those intubated first. However, those same patients also experience a greater hazard of being discharged alive up to about 15-35 days, after which that hazard decreases either to the point of reversing direction (for NIRS in general and HFNO specifically) or no longer being statistically significant (for NIPPV). Essentially, patients initially treated noninvasively have higher hazards of both in-hospital death and discharge alive up to about a month. After that, their hazard of in-hospital death remains elevated but their hazard of discharge alive decreases. Moreover, those started on HFNO first experience faster intubation compared to those started on NIPPV first.

These results offer several important contributions. Studies utilizing NIRS for patients with COVID-19 compare one or both noninvasive respiratory support strategies either to each other or to conventional oxygen therapy and have reported mixed findings. Some report improved outcomes with high flow nasal oxygen compared to conventional oxygen therapy,^41–43^ or to noninvasive positive pressure ventilation,^44^ while others report improved outcomes with noninvasive positive pressure ventilation compared to conventional oxygen therapy,^45^ or to high flow nasal oxygen.^46^ Yet some studies report no difference in outcomes compared to conventional oxygen therapy,^43, 45, 47^ or to each other.^46–50^ Those studies utilize intubation directly as the primary outcome alone or as part of a composite outcome with mortality.^>41–>48, >51, >52^ We found that invasive mechanical ventilation as a comparator intervention may be the optimal method of respiratory support for COVID-19 patients that need more than conventional oxygen.

Additionally, most studies are limited to patients admitted to the ICU, which carries inherent selection bias and confounding potentially reflected as higher intubation rates of around 50%. We included all patients regardless of location as many patients were cared for outside of the physical ICU location or the ICU service from pandemic-strained resources.

We found that the benefit of leaving the hospital alive has a relationship with time. There are several potential explanations for this. NIRS cohorts are classified by a phenotyping algorithm based on the first therapy received but could have crossed over between NIRS modalities at any point with or without intubation. Crossover in those patients requires failing NIRS twice before intubation. Second, intubation could have been delayed for various reasons at the cost of worsening acute lung injury that may have benefitted from lung protective ventilation. Gershengorn found similar results in that the overall 46% failure rate on HFNO evolved from a decreased odds of failure for those on HFNO for a short time to an increased odds of failure for those on HFNO for longer.^52^ Similar findings were reported with NIPPV, which reduced mortality only in patients with short hospital stays, but was associated with higher mortality in those hospitalized longer than seven days.^53^

There are several issues to consider when generalizing and interpreting data on noninvasive respiratory support and invasive mechanical ventilation in COVID-19 patients. Early in the pandemic, early intubation was often recommended to avoid aerosol exposure to healthcare staff and based on the high failure rates of NIPPV in the SARS epidemic. This was followed by a concerningly high early mortality reported in patients on mechanical ventilation^22^ and debate over the clinical syndrome seen in COVID-19 that contributed to variability in mechanical ventilation (e.g., high tidal volumes or alternative ventilator modes) or an avoidance of intubation altogether in some patients.^54–59^ Simultaneously, pharmacological adjuncts to standard of care have changed over time, especially during the duration of this study^60^, including convalescent plasma ^61–63^, corticosteroids,^64–68^ interleukin ^69–76^ and janus kinase ^77–79^ inhibitors, antivirals ^80–83^, hydroxychloroquine ^80, 84, 85^, and anticoagulation strategies.^86–88^ Lastly, patient surges^89^ and surge capacity almost certainly contributed to patient outcomes as more patients were managed outside of traditional ICUs, or in expansion ICUs, and facilities were faced with impending ventilator shortages, staffing ratio changes, and an increase in traveling staff. These results are, however, hypothesis generating for non-COVID-19 acute respiratory distress syndrome patients where NIPPV has been shown in some studies to be associated with higher mortality than IMV.^90^

There are also important limitations to our study. Our data are limited to the COVID-19 patients in 2020 and the evolution of COVID-19 management potentially limits generalizability. Additionally, results are based on the first assigned therapy and patients who crossed over between NIRS and any imbalance between those crossovers could confound the findings. There were also some necessary assumptions based on the nature of the observational dataset from the electronic health record. For instance, we only considered a given patient’s first COVID-19 hospital visit where they required respiratory support. If two visits that appeared to belong to the same person were less than 24 hours apart, we assumed a hospital transfer occurred and combined these records into the same visit. We attempted to control for confounding, including by indication, by weighting our analysis by the inverse probability for treatment assignment, further adjusting for potential confounders in the Cox models, and by a competing risks analysis where death and intubation were competing risks. Additionally, some patients had evidence of all three treatments but no clear treatment ordering for which we performed sensitivity analyses to assess how dependent our results are on our specific inclusion, exclusion, or imputation decisions. Lastly, clinical care was not protocolized and there were likely important differences between flow rates with HFNO, airway pressures with NIPPV (of which only facemask NIPPV was available), and mechanical ventilator settings that could confound these results. Despite these limitations, the size of our dataset and statistical analyses still provide valuable results.

We found that patients intubated early without a trial of noninvasive respiratory support had a lower hazard of in-hospital death, but also a lower hazard of hospital discharge alive up to about a month. Clinical focus should shift to not delaying intubation in patients in whom noninvasive respiratory support has not reduced work of breathing or hypoxemia. Meanwhile, studies are needed to identify optimal patients for each noninvasive respiratory support modality and early prediction and identification of failure of that modality.

## Statements and Declarations

### Author Contributions

JMM, VS, JMF, and PE conceived the study idea. PE, VS, and JMM developed the phenotyping algorithm. PE, JMF, JMM, and SP preprocessed the data. PE and VS applied the phenotyping algorithm. JMF, EJB, and SP performed the statistical analysis. JMM and JMF drafted the initial manuscript, and all authors participated in manuscript revisions.

## Data Availability

Data are available upon reasonable request, IRB approval, and a data use agreement.

## Acknowledgements

The authors would like to thank Don Saner and Mario Arteaga from the Banner Health Network Clinical Data Warehouse for their support during this project.

## Online Supplement

### Data Preprocessing

Data were preprocessed to exclude participants under age 18, visits for which a phenotype was not available, and participants who exited the hospital (either alive or dead) before treatment. Some patients appeared to have multiple applicable hospital visits. Hospital transfers were identified by looking for multiple entries with the same ID where the patients had the same gender, race, ethnicity, age (max one year difference), the patient was alive at transfer, and consecutive hospital discharge and admission times were fewer than 24 hours apart. Identified transfers were combined into a single visit entry, and for six visits, phenotypes were adjusted as appropriate. Overall, 48 transfers were identified, and 122 later visits were discarded. Later visits were discarded in order to sample from the population of first COVID-19 visits. Height and weight were defined preferentially as the first recorded height and weight and secondarily as the admission height and weight (available for ICU data only); among ICU visits, there were 72 instances of using an admission instead of a time-stamped height and 11 instances of using an admission instead of a time-stamped weight. Heights below 48 inches and weights below 50 pounds were classified as potential data errors, and the next valid height and/or weight was sought. For nine people, this resulted in no height data being available; for two people these rules resulted in no weight data being available. In the ICU data, 72 visits had an admission height but not a time-stamped height, and 11 visits had an admission weight but not a time-stamped weight. In order to work with a deidentified data set, for the 42 participants with age over 89 years (18 participants with “>89”, and 24 with specific ages >89), their age was marked as 90. Race and ethnicity were standardized. For the respiratory rate, FiO_2_, oxygen flow rate, and SpO_2_ variables, the value closest to and before first treatment were identified. For people with phenotype 8 (evidence of all three treatments but unclear order), the first treatment start time was taken to be the earliest treatment start time identified by the phenotyping algorithm. Oxygen flow rates were used to calculate FiO_2_ (FiO_2_ = 100(0.21 + oxygen flow [L/min^-1^] x 0.03), and the FiO_2_ value (either raw or calculated) selected for later use was the latter one.

### Propensity Score Estimation via Generalized Boosted Models

We follow McCaffrey et al.’s (2013) approach to estimating propensity scores by using generalized boosted models (GBMs).^1^ GBMs estimate propensity scores for the three treatments by fitting the models to the dichotomized outcomes of each treatment versus the other two. This results in three GBMs being fit each time propensity scores are estimated for models comparing NIPPV, HFNO, and IMV. Because each GBM consists of an iterative process of combining simple regression trees into a piecewise constant function, for each GBM, we must choose a point at which to stop adding onto the regression tree. Four criteria can readily be used to choose a stopping point; all four attempt to describe the difference between the distributions of covariates for the comparison groups for each dichotomized outcome (e.g., between patients who received IMV and all patients); the goal is to minimize the differences between the groups. Note that we wish to estimate the average treatment effect between pairs of treatments within the context of our entire population. This is considered in each stopping criterion:^1^

1. *Mean Absolute Standardized Bias*: mean across covariates of the absolute standardized difference between the weighted mean of the covariate for the treatment group of interest and the unweighted mean of the covariate for the entire sample. Weights are the inverse of the estimated propensity scores. Because we are interested in estimating the treatment effect in the general population of people receiving any of the three treatments, we standardize (i.e., divide) here by the estimated standard deviation from the unweighted, pooled, full sample.
2. *Maximum Absolute Standardized Bias*: maximum across covariates of the absolute standardized difference between the weighted mean of the covariate for the treatment group of interest and the unweighted mean of the covariate for the entire sample. Weights and standardization are as described above.
3. *Mean Kolmogorov-Smirnov* mean across covariates of the supremum of pointwise absolute differences between the weighted empirical distribution function for the covariate among those treated with the treatment of interest and the unweighted empirical distribution function for the entire sample.
4. *Maximum Kolmogorov-Smirnov*: maximum across covariates of the supremum of pointwise absolute differences between the weighted empirical distribution function for the covariate among those treated with the treatment of interest and the unweighted empirical distribution function for the entire sample.

For each GBM of a dichotomized outcome, we allow a maximum of 5000 simple regression trees. Empirically, this number appeared to allow all four criteria above to achieve their minimum and thus was large enough without being excessively large. For each outcome and each stopping criterion, the optimal number of trees was selected. Thus, for each treatment, we estimated four propensity scores for having been given the treatment in question. Inverse probability of treatment weighting was used in subsequent analyses by taking the inverse of the propensity scores.

For all non-interaction and non-sensitivity models, a separate analysis was done for each stopping criterion to ensure that results were not dependent on the particular criterion used. No result was dependent on stopping criterion.

### Sensitivity Analyses for Time to Intubation

The competing risks analyses of time-to-intubation made a series of analytical decisions that could impact the subsequent inference. We thus designed a series of sensitivity analyses that systematically varied our assumptions and analytical approach to ensure that results were not dependent on specific decisions.

The primary reason for sensitivity analyses was that 326 visits could not be reliably classified using the phenotyping algorithm. These unclassifiable visits always contained evidence of all three treatments (IMV, HFNO, and NIPPV). However, in these cases, conflicting information in the medical record often made it unclear to the algorithm which treatment came first. For example, a patient might appear to be intubated, switched to NIPPV, then reintubated in a 30-minute period. Such information would cause the algorithm to fail to assign a treatment ordering to that visit. While these same cases were present in the other analyses, in those analyses, we were able to impute treatment, estimate propensity scores for each imputed data set, model each data set, then combine model estimates across data sets using Rubin’s Rules.

The weighted Gray’s test of competing risks, however, used a bootstrapped null distribution. There was thus no obvious way to use Rubin’s Rules. A common alternative approach to combining propensity scores and multiple imputation --- estimating propensity scores for each imputed data set, averaging them, and choosing a single imputed data set at random to use in the analysis along with the averaged propensity scores --- wasn’t applicable because of the need to impute treatment. When treatment is imputed, it is unclear how well a particular data set approximates the true but unknown treatment identities, and thus using the average propensity score approach is inappropriate.^2^ Moreover, different observations could be included in each data set since people imputed to have an invasive first treatment would be excluded. Determining how to compute average propensity scores for people who were not consistently included in the analyses and how to appropriately handle the data from those individuals left out of the randomly sampled data set is beyond the scope of this paper. A final challenge of using the average propensity score approach came from the need to impute the time to event for 52 people; if the imputed outcomes were particularly outlying and influential for a randomly selected data set, then the inference may not be correct. An alternative to Rubin’s Rules — judging statistical significance via the median p-value from the pooled p-values across all the imputed data sets, while appealing, is less common and would need to be utilized in an analysis scenario not covered in Eekhout et al.^3^

The sensitivity analyses thus encompass a range of ways to handle the above challenges. Each combines a data decision, an outcome decision, an imputation decision, and an inference approach decision to arrive at an analysis pipeline. Note that some combinations of the four decisions were not possible. Moreover, where it was possible to take either the average propensity score or median p-value approach, we did not always conduct both corresponding analyses. Rather, we defaulted to the average propensity score approach except when we are imputing time-to-event information; in those cases, we also conduct the corresponding analysis with the median p-value inference approach as an extra confirmation that the specific data set chosen in the average propensity score approach didn’t have particularly influential imputed times-to-event.

**Data Decision:** How and how much of the unclassifiable data to include.

- Option A: We assume that we know everything about the unclassifiable data and base our knowledge on treatment start information. We exclude patients with IMV as the first marked treatment and assign patients with HFNO or NIPPV marked as the first treatment to the corresponding treatment group. If the two noninvasive therapies tie for first treatment (7 people), we randomly select one.

- Option B: We assume that we know whether the first treatment was IMV or NIRS based on the treatment time stamps. Patients with IMV as the first treatment are excluded from analysis. However, because of potential nomenclature inconsistencies between NIPPV and HFNO, we assume that the first noninvasive treatment could have been either NIPPV or HFNO. We impute the initial noninvasive treatment type.

- Option C: We assume that we do not know what treatment came first and thus whether or not anyone with the unclassifiable phenotype should be included in this competing risks analysis. Thus, we exclude them all from the analysis.

**Unknown Outcome Decision:**

- Yes: Include patients with unknown time-to-event.
- No: Do not include patients with unknown time-to-event.

**Imputation Decision:** The decision whether to do imputation impacts which predictors are used in the propensity score calculations. If imputation is conducted, the propensity score algorithm uses the previously described predictors; otherwise, it uses a restricted subset for which all information is available (age, gender, hospital size).

- Yes: Conduct multiple imputation by chained equations.
- No: Do not impute missing data.

**Inference Approach Decision:**

- Average Propensity Score Approach: In this approach, the propensity scores for each visit are averaged across imputed data sets. These averaged scores are used with a randomly-selected imputation dataset. This approach is not appropriate when treatment is imputed.
- Median P-Value Approach: In this approach, the weighted Gray’s test is conducted on each imputed data set (with its own set of propensity scores). Then, the median p-value across data sets is taken for inference.
- Single Propensity Score Approach: This case arises when we are not imputing data. Then, we estimate propensity scores once (based on the limited predictor set) and use them in the weighted Gray’s test.

The ten proposed competing risks analyses correspond to the following combinations of the four decision points (all p-values were <0.001):

| Analysis | Data Decision | Unknown Outcome Decision | Imputation Decision | Inference Approach Decision |
| --- | --- | --- | --- | --- |
| Sensitivity Analysis #1 | A | Yes | Yes | Average Propensity Score |
| Sensitivity Analysis #2 | A | Yes | Yes | Median P-Value |
| Sensitivity Analysis #3 | A | No | Yes | Average Propensity Score |
| Sensitivity Analysis #4 | A | No | No | Single Propensity Score |
| Sensitivity Analysis #5 | B | Yes | Yes | Median P-Value |
| **Primary Competing Risks Analysis** | **B** | **No** | **Yes** | **Median P-Value** |
| Sensitivity Analysis #6 | C | Yes | Yes | Average Propensity Score |
| Sensitivity Analysis #7 | C | Yes | Yes | Median P-Value |
| Sensitivity Analysis #8 | C | No | Yes | Average Propensity Score |
| Sensitivity Analysis #9 | C | No | No | Single Propensity Score |

We considered the analysis Data Decision B + Unknown Outcome Decision: No + Imputation Decision: Yes + Inference Approach Decision: Median P-Value to be primary and the others to be sensitivity analyses. Additionally, given the lack of inferential difference associated with the different propensity score estimation stopping rules, for all sensitivity analyses, we only used the mean absolute standardized bias stopping rule. We did, however, use all four stopping rules for the primary competing risks analysis.

**Figure 1:**
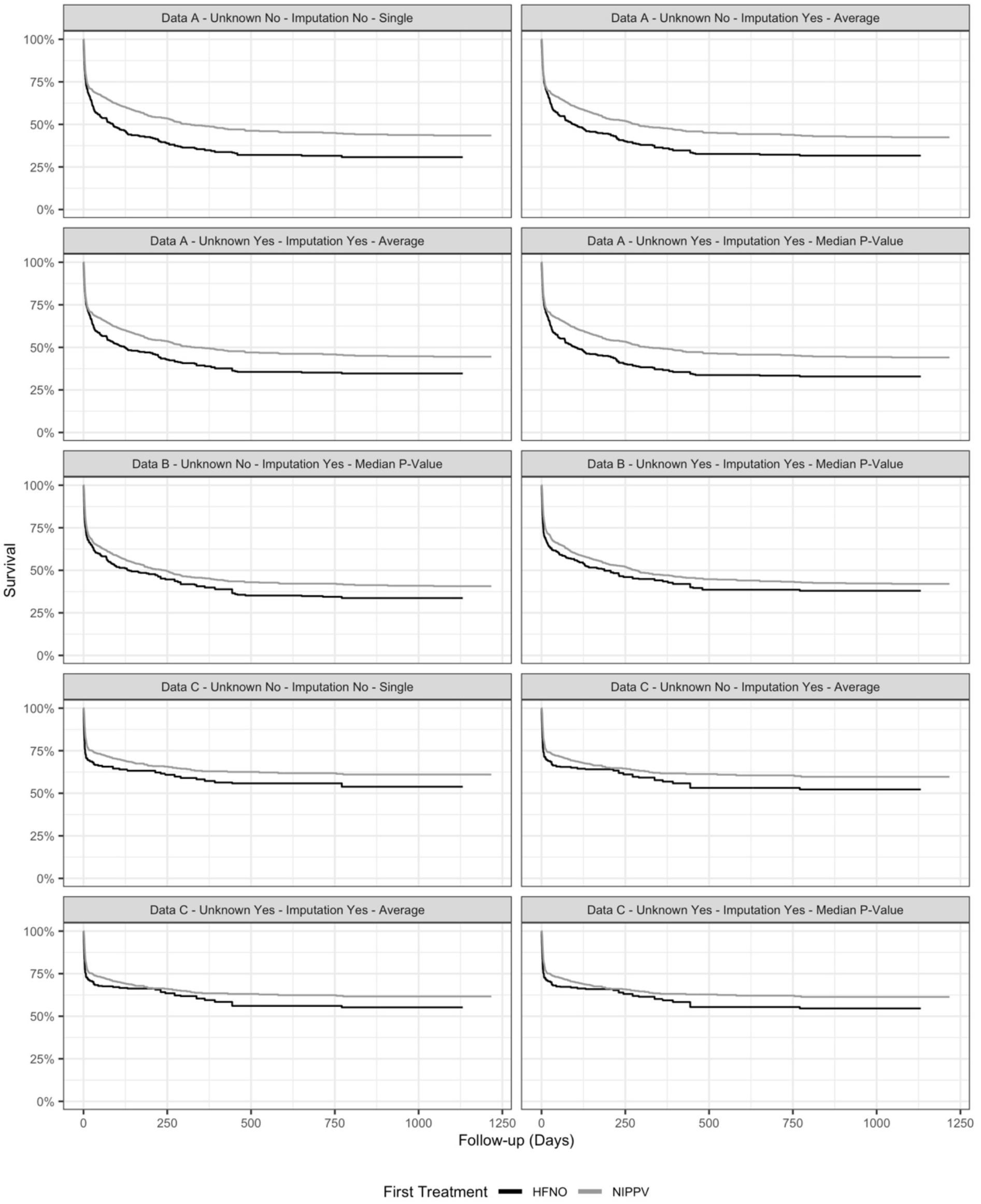
Time-to-intubation sensitivity analyses. In the estimation of these weighted survival plots, people who die remain in the risk set after their death because they have neither been censored nor experienced intubation. All time-to-intubation analyses showed statistically significant differences between initial treatment with **HFNO** and NIPPV (p < 0.001).

**Table 1:** Demographics of Unclassifiable Cohort

| Measure | Unknown |
| --- | --- |
| N (%) | 326 (14% of 2354) |
| Sex |  |
| Male | 222 (68% of 326) |
| Female | 104 (32% of 326) |
| Age | 65 (58 - 71) |
| BMI | 31.82 (28.06 - 37.66) |
| Ethnicity |  |
| Not Hispanic or Latino | 165 (53% of 314) |
| Hispanic or Latino | 149 (47% of 314) |
| Race |  |
| White | 245 (78% of 313) |
| Black or African American | 16 (5% of 313) |
| Asian/Native Hawaiian/Other Pacific Islander | 5 (2% of 313) |
| American Indian or Alaska Native | 30 (10% of 313) |
| Other | 17 (5% of 313) |
| APACHE IVa Score on Admission | 65 (51.75 - 82) |
| Vital Signs on Treatment Assignment |  |
| HR | 87 (74 - 98.75) |
| SBP | 129 (114 - 144.5) |
| DBP | 74 (65 - 82) |
| SpO2 | 91 (88 - 94) |
| Oxygen Flow Rate/L | 40 (15 - 50) |
| FiO2* | 100 (80 - 100) |
| SpO2:FiO2 | 95 (90 - 119.22) |
| Temperature (C) | 37 (37 - 37) |
| Respiratory Rate | 25 (20 - 31.25) |
| Comorbidities** |  |
| Diabetes | 203 (68% of 299) |
| Chronic Kidney Disease | 61 (20% of 299) |
| Heart Failure | 50 (17% of 299) |
| Hypertension | 246 (82% of 299) |
| Chronic Liver Disease | 74 (25% of 299) |
| Neoplasm or Immunosuppression | 21 (7% of 299) |
| COPD | 81 (27% of 299) |
| Labs on Admission |  |
| PaO2 (Worst Value) | 62.55 (55.73 - 73) |
| PaO2:FiO2 (Worst Value) | 66.33 (56 - 90.32) |
| White Blood Cell Count | 8 (6 - 11.4) |
| Lactate | 1.6 (1.2 - 2.38) |
| pH | 7.42 (7.36 - 7.45) |
| PaCO2 | 35 (30.6 - 41) |
| HCO3 | 22 (20 - 24) |
| BNP | 356 (140 - 1184) |
| Creatinine | 0.97 (0.75 - 1.31) |
| Therapies |  |
| Therapeutic Anticoagulation<sup>a</sup> | 307 (94% of 326) |
| Hydroxychloroquine | 12 (4% of 326) |
| Remdesivir | 237 (73% of 326) |
| Corticosteroids<sup>b</sup> | 318 (98% of 326) |
| Tocilizumab | 0 (0% of 326) |
| Continuous Paralysis | 132 (40% of 326) |
| Time from Hospital Admission to First Treatment Assignment (Hours) | 114.32 (38.65 - 238.4) |
| Vasopressor Use |  |
| Before Treatment | 32 (10% of 326) |
| At or After Treatment | 307 (94% of 326) |
| Location at Treatment Assignment |  |
| Emergency Department | 22 (7% of 326) |
| ICU | 145 (44% of 326) |
| Ward | 53 (16% of 326) |
| Stepdown | 106 (33% of 326) |

**Table 2:**
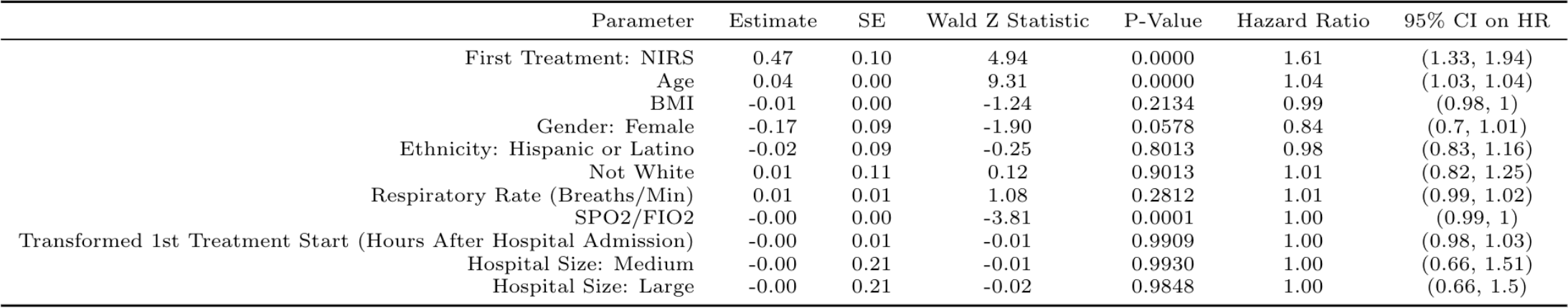
Time-to-In-Hospital Death (NIRS vs IMV). Model estimates shown below correspond to the mean absolute standardized bias stopping rule in the generalized boosted models. Inferential results were the same for the other stopping rules

**Figure 2:**
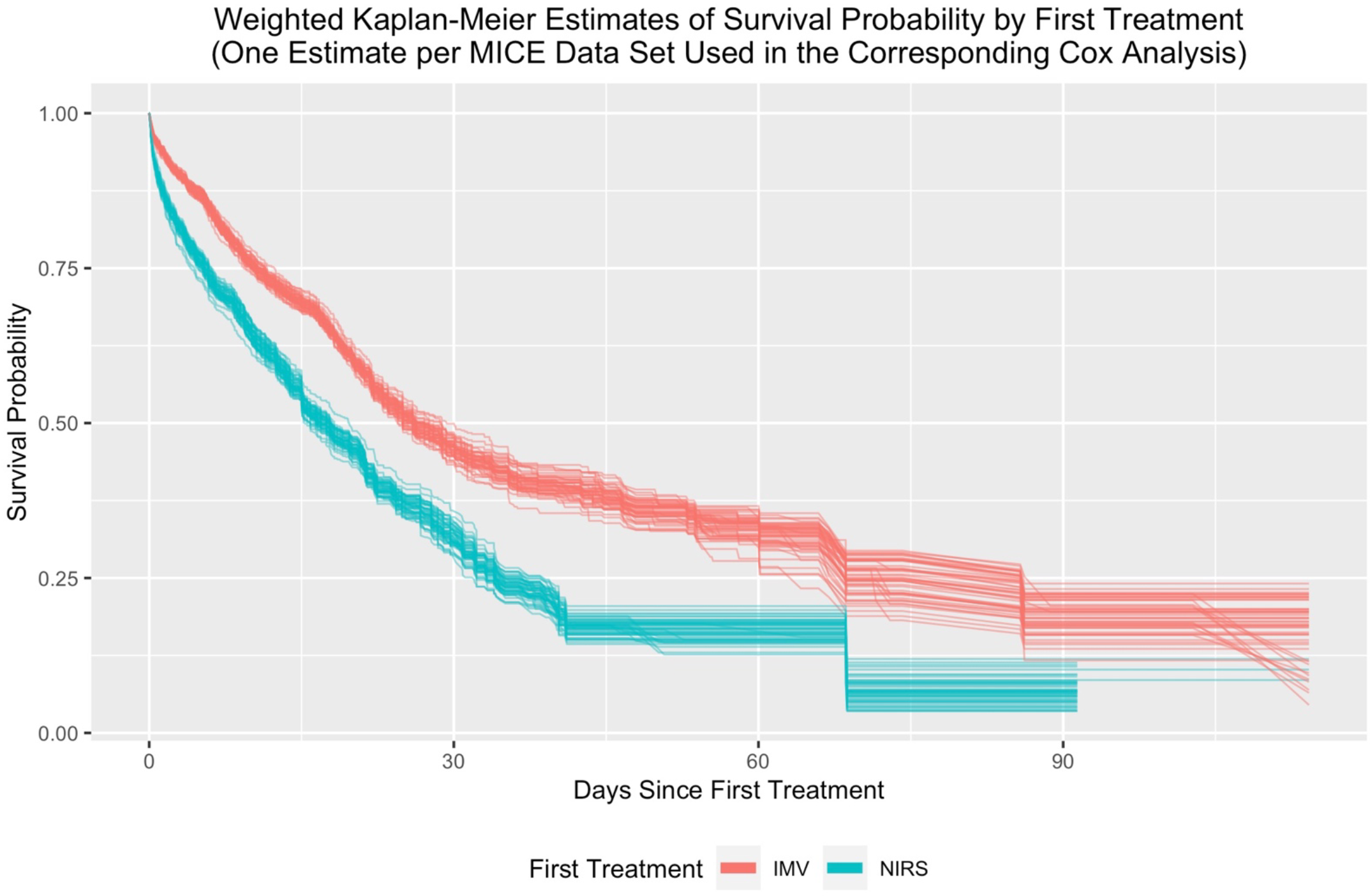
Time-to-In-Hospital Death (NIRS vs IMV). Time-to-in-hospital death weighted Kaplan-Meier curves for noninvasive respiratory support versus invasive mechanical ventilation. Curves use propensity scores estimated via the mean absolute standardized bias stopping rule in the generalized boosted models.

**Table 3:** Time-to-Hospital Discharge Alive (NIRS vs IMV). Death is treated as a competing risk. Time zero is time of first treatment assignment.

| Parameter | Estimate | SE | Wald Z Statistic | P-Value | Hazard Ratio | 95% CI on HR |
| --- | --- | --- | --- | --- | --- | --- |
| First Treatment: NIRS | 1.09 | 0.12 | 9.05 | 0.0000 | 2.98 | (2.35, 3.78) |
| Time x NIRS | -0.03 | 0.01 | -4.67 | 0.0000 | 0.97 | (0.96, 0.98) |
| Age | -0.00 | 0.00 | -1.43 | 0.1519 | 1.00 | (0.99, 1) |
| BMI | 0.01 | 0.00 | 2.61 | 0.0090 | 1.01 | (1, 1.01) |
| Gender: Female | 0.23 | 0.07 | 3.25 | 0.0011 | 1.26 | (1.1, 1.45) |
| Ethnicity: Hispanic or Latino | -0.15 | 0.08 | -1.91 | 0.0556 | 0.86 | (0.74, 1) |
| Not White | -0.32 | 0.09 | -3.74 | 0.0002 | 0.72 | (0.61, 0.86) |
| Respiratory Rate (Breaths/Min) | -0.02 | 0.01 | -3.02 | 0.0025 | 0.98 | (0.97, 0.99) |
| SPO2/FIO2 | 0.01 | 0.00 | 10.11 | 0.0000 | 1.01 | (1, 1.01) |
| Transformed 1st Treatment Start (Hours After Hospital Admission) | 0.01 | 0.01 | 0.82 | 0.4107 | 1.01 | (0.99, 1.02) |
| Hospital Size: Medium | -0.71 | 0.18 | -3.97 | 0.0001 | 0.49 | (0.35, 0.7) |
| Hospital Size: Large | -0.93 | 0.18 | -5.24 | 0.0000 | 0.39 | (0.28, 0.56) |

**Figure 3:**
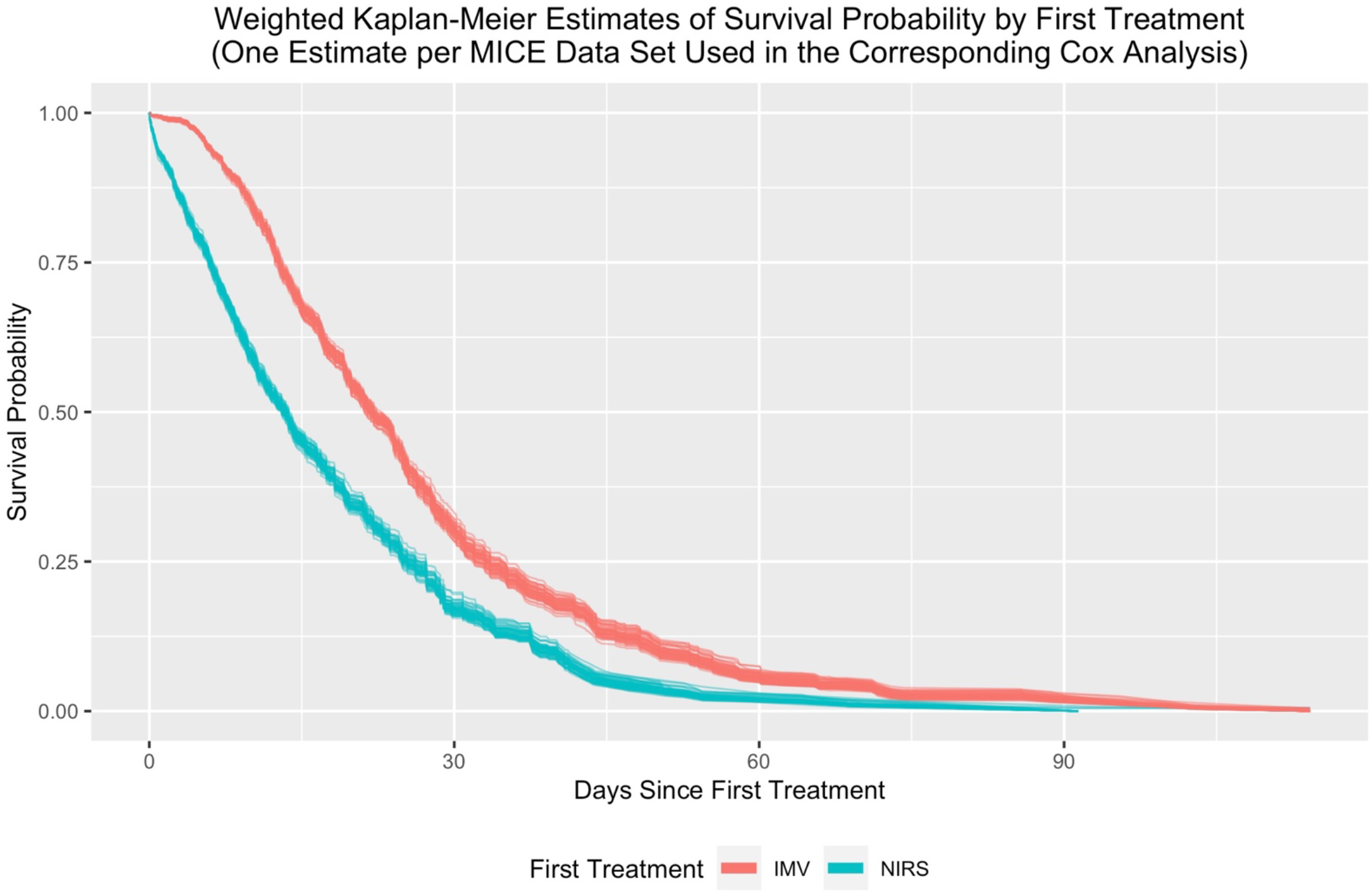
Time-to-Hospital Discharge Alive (NIRS vs IMV). Time-to-hospital discharge alive weighted Kaplan-Meier curves for noninvasive respiratory support versus invasive mechanical ventilation. Time zero is time of first treatment assignment. In these Kaplan-Meier estimates, people experiencing death are treated as censored. Curves use propensity scores estimated via the mean absolute standardized bias stopping rule in the generalized boosted models.

**Table 4:** Time-to-Hospital Discharge Alive (NIRS vs IMV) Sensitivity Analysis. Death is treated as a competing risk. Time zero is time of hospital admission.

| Parameter | Estimate | SE | Wald Z Statistic | P-Value | Hazard Ratio | 95% CI on HR |
| --- | --- | --- | --- | --- | --- | --- |
| First Treatment: NIRS | 0.90 | 0.13 | 7.01 | 0.0000 | 2.47 | (1.92, 3.18) |
| Time x NIRS | -0.02 | 0.01 | -3.14 | 0.0017 | 0.98 | (0.97, 0.99) |
| Age | -0.00 | 0.00 | -1.59 | 0.1122 | 1.00 | (0.99, 1) |
| BMI | 0.01 | 0.00 | 2.76 | 0.0057 | 1.01 | (1, 1.02) |
| Gender: Female | 0.23 | 0.07 | 3.24 | 0.0012 | 1.26 | (1.09, 1.44) |
| Ethnicity: Hispanic or Latino | -0.19 | 0.08 | -2.45 | 0.0142 | 0.83 | (0.72, 0.96) |
| Not White | -0.26 | 0.09 | -2.99 | 0.0028 | 0.77 | (0.65, 0.91) |
| Respiratory Rate (Breaths/Min) | -0.02 | 0.01 | -3.67 | 0.0002 | 0.98 | (0.97, 0.99) |
| SPO2/FIO2 | 0.01 | 0.00 | 11.36 | 0.0000 | 1.01 | (1, 1.01) |
| Transformed 1st Treatment Start (Hours After Hospital Admission) | -0.09 | 0.01 | -9.83 | 0.0000 | 0.92 | (0.9, 0.93) |
| Hospital Size: Medium | -0.61 | 0.18 | -3.42 | 0.0006 | 0.54 | (0.38, 0.77) |
| Hospital Size: Large | -0.78 | 0.18 | -4.40 | 0.0000 | 0.46 | (0.33, 0.65) |

**Figure 4:**
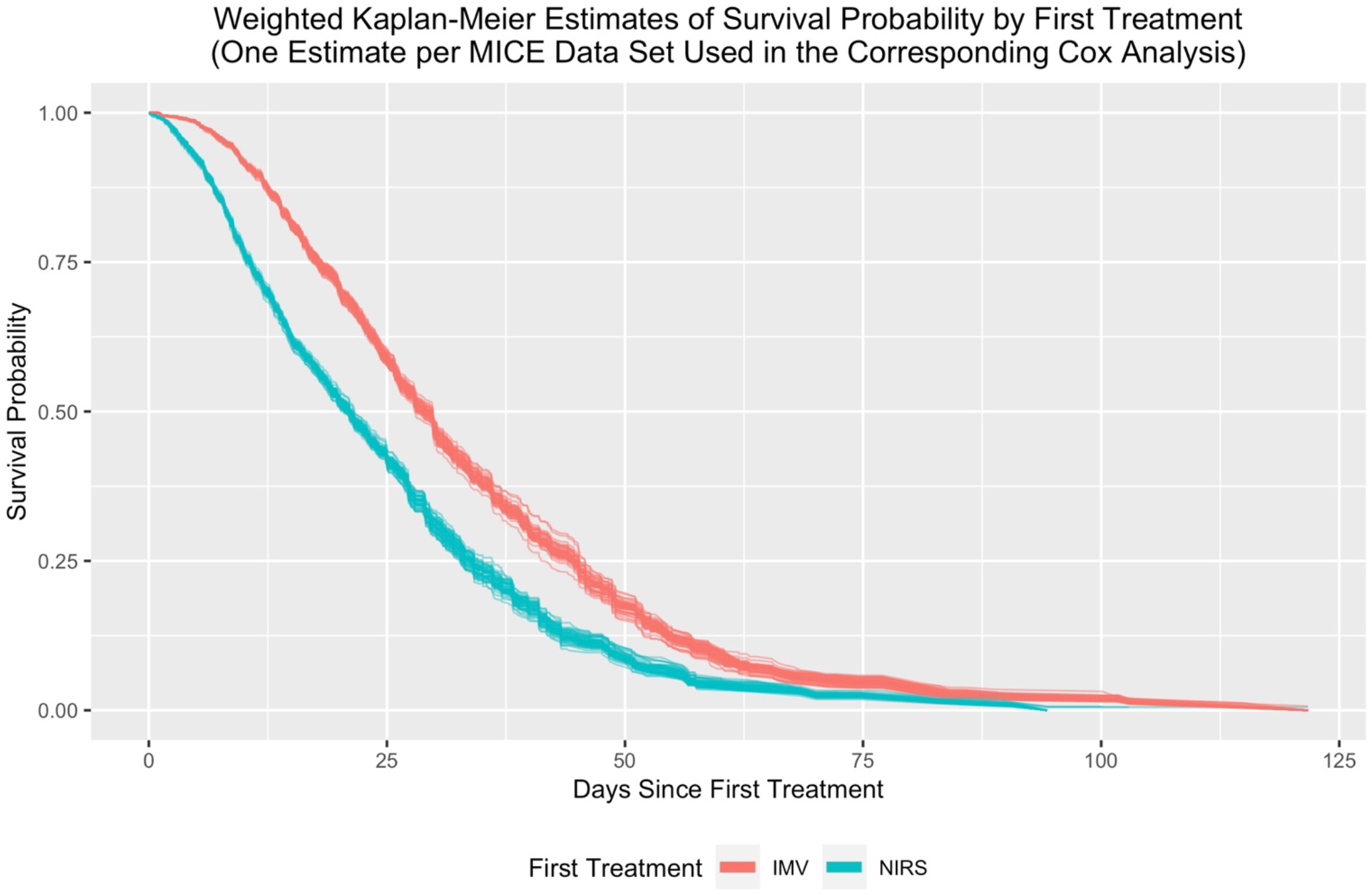
Time-to-Hospital Discharge Alive (NIRS vs IMV) Sensitivity Analysis. Time-to-hospital discharge alive weighted Kaplan-Meier curves for noninvasive respiratory support versus invasive mechanical ventilation. Time zero is time of hospital admission. In these Kaplan-Meier estimates, people experiencing death are treated as censored. Curves use propensity scores estimated via the mean absolute standardized bias stopping rule in the generalized boosted models.

**Table 5:** Time-to-ICU Discharge Alive (NIRS vs IMV), with death as a competing risk for noninvasive respiratory support versus invasive mechanical ventilation. Time zero is time of ICU admission.

| Parameter | Estimate | SE | Wald Z Statistic | P-Value | Hazard Ratio | 95% CI on HR |
| --- | --- | --- | --- | --- | --- | --- |
| First Treatment: NIRS | 0.29 | 0.12 | 2.37 | 0.0178 | 1.33 | (1.05, 1.69) |
| Time x NIRS | -0.02 | 0.01 | -2.14 | 0.0321 | 0.98 | (0.97, 1) |
| Age | -0.00 | 0.00 | -0.96 | 0.3357 | 1.00 | (0.99, 1) |
| BMI | 0.00 | 0.00 | 0.49 | 0.6209 | 1.00 | (0.99, 1.01) |
| Gender: Female | 0.10 | 0.08 | 1.32 | 0.1859 | 1.11 | (0.95, 1.28) |
| Ethnicity: Hispanic or Latino | -0.03 | 0.08 | -0.36 | 0.7210 | 0.97 | (0.83, 1.14) |
| Not White | -0.04 | 0.09 | -0.49 | 0.6240 | 0.96 | (0.8, 1.14) |
| Respiratory Rate (Breaths/Min) | -0.01 | 0.01 | -2.28 | 0.0227 | 0.99 | (0.98, 1) |
| SPO2/FIO2 | 0.01 | 0.00 | 7.35 | 0.0000 | 1.01 | (1, 1.01) |
| Transformed 1st Treatment Start (Hours After ICU Admission) | -0.07 | 0.01 | -5.44 | 0.0000 | 0.94 | (0.92, 0.96) |
| Hospital Size: Medium | -0.53 | 0.18 | -2.94 | 0.0033 | 0.59 | (0.42, 0.84) |
| Hospital Size: Large | -0.56 | 0.17 | -3.25 | 0.0011 | 0.57 | (0.41, 0.8) |

**Figure 5:**
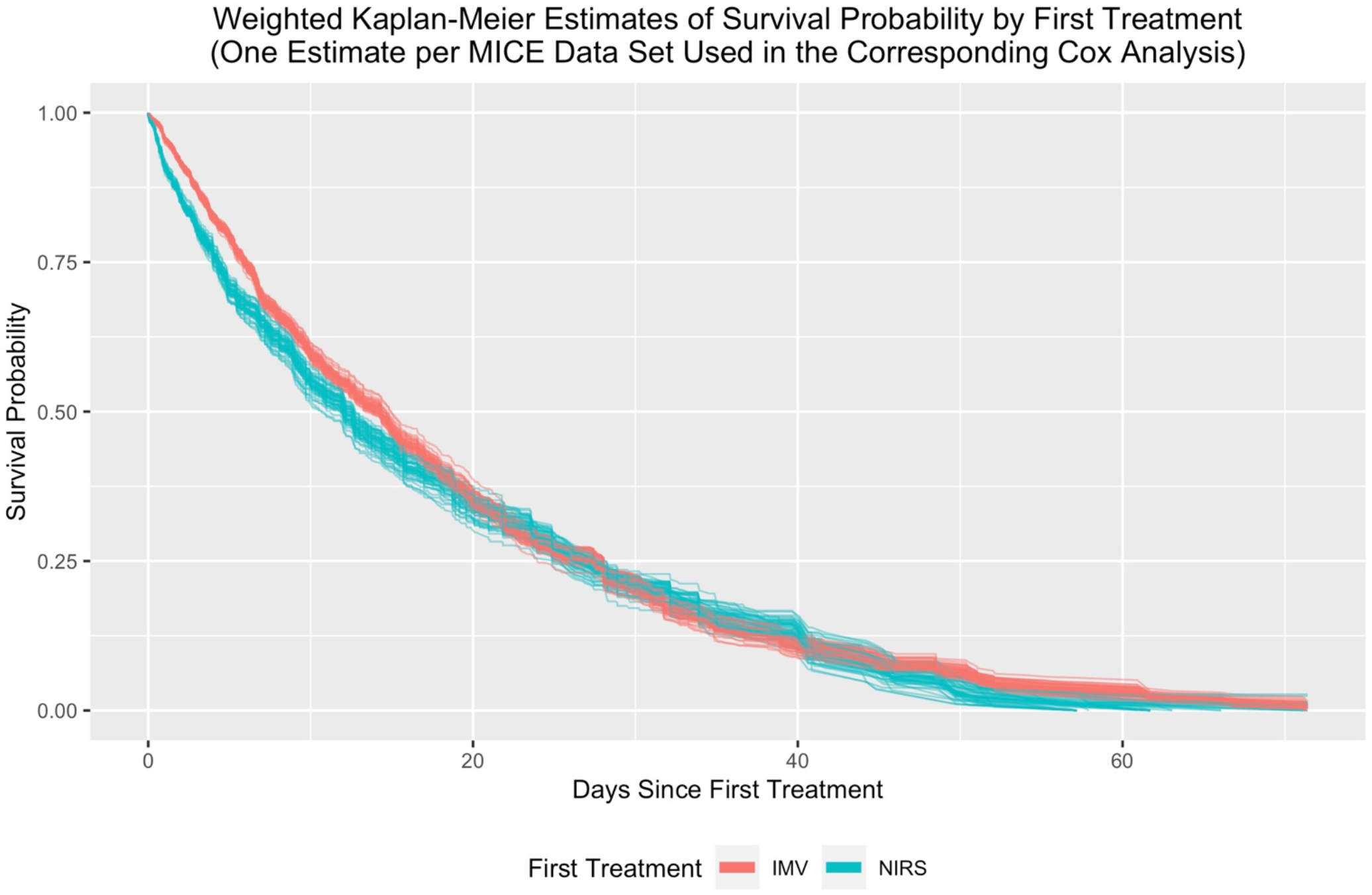
Time-to-ICU Discharge Alive Secondary Outcome (NIRS vs IMV). Weighted Kaplan-Meier curves for noninvasive respiratory support versus invasive mechanical ventilation. Time zero is time of ICU admission. In these Kaplan-Meier estimates, people experiencing death are treated as censored. Curves use propensity scores estimated via the mean absolute standardized bias stopping rule in the generalized boosted models.

**Table 6:** Time-to-In-Hospital Death (HFNO vs NIPPV vs IMV). Model estimates shown below correspond to the mean absolute standardized bias stopping rule in the generalized boosted models. Inferential results were the same for the other stopping rules.

| Parameter | Estimate | SE | Wald Z Statistic | P-Value | Hazard Ratio | 95% CI on HR |
| --- | --- | --- | --- | --- | --- | --- |
| First Treatment: HFNO | 0.86 | 0.13 | 6.82 | 0.0000 | 2.37 | (1.85, 3.03) |
| First Treatment: NIPPV | 0.36 | 0.10 | 3.66 | 0.0003 | 1.44 | (1.18, 1.74) |
| Age | 0.04 | 0.00 | 9.40 | 0.0000 | 1.04 | (1.03, 1.05) |
| BMI | -0.00 | 0.01 | -0.71 | 0.4798 | 1.00 | (0.98, 1.01) |
| Gender: Female | -0.21 | 0.10 | -2.05 | 0.0406 | 0.81 | (0.66, 0.99) |
| Ethnicity: Hispanic or Latino | -0.07 | 0.10 | -0.72 | 0.4709 | 0.93 | (0.77, 1.13) |
| Not White | 0.08 | 0.11 | 0.75 | 0.4546 | 1.09 | (0.87, 1.35) |
| Respiratory Rate (Breaths/Min) | 0.01 | 0.01 | 1.36 | 0.1745 | 1.01 | (1, 1.02) |
| SPO2/FIO2 | -0.00 | 0.00 | -3.36 | 0.0008 | 1.00 | (0.99, 1) |
| Transformed 1st Treatment Start (Hours After Hospital Admission) | 0.00 | 0.01 | 0.31 | 0.7565 | 1.00 | (0.98, 1.03) |
| Hospital Size: Medium | 0.16 | 0.24 | 0.66 | 0.5105 | 1.17 | (0.73, 1.89) |
| Hospital Size: Large | 0.09 | 0.24 | 0.38 | 0.7023 | 1.10 | (0.68, 1.77) |
| HFNO - NIPPV | 0.50 | 0.12 | 4.03 | 0.0001 | 1.65 | (1.29, 2.1) |

**Figure 6:**
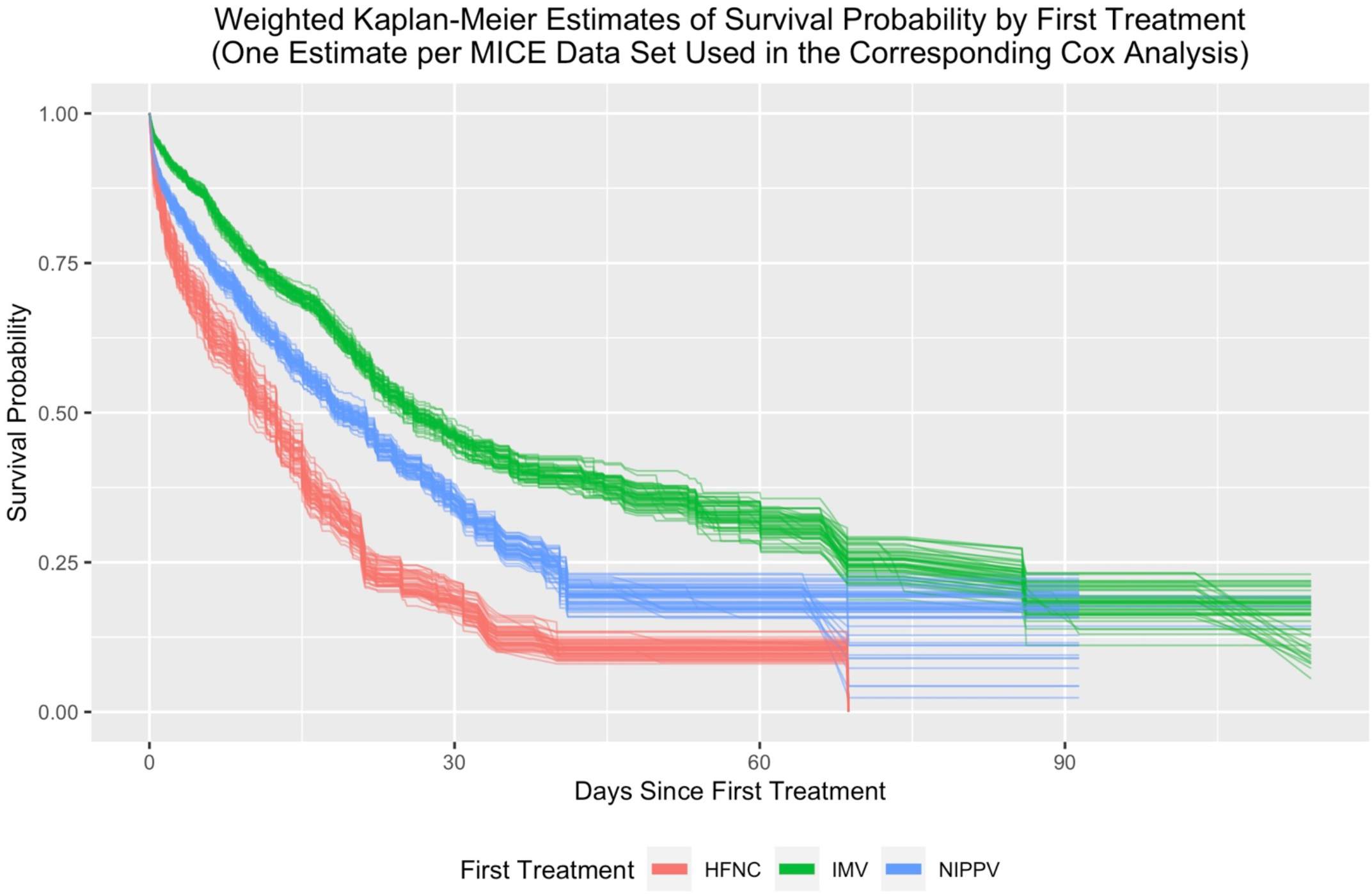
Time-to-In-Hospital Death (HFNO vs NIPPV vs IMV). Time-to-in-hospital death weighted Kaplan-Meier curves for HFNO versus NIPPV versus invasive mechanical ventilation. Curves use propensity scores estimated via the mean absolute standardized bias stopping rule in the generalized boosted models.

**Table 7:** Time-to-Hospital Discharge Alive (HFNO vs NIPPV vs IMV). Death is treated as a competing risk. Time zero is time of first treatment assignment.

|  | Parameter | Estimate | SE | Wald Z Statistic | P-Value | Hazard Ratio | 95% CI on HR |
| --- | --- | --- | --- | --- | --- | --- | --- |
|  | First Treatment: HFNO | 0.97 | 0.24 | 4.04 | 0.0001 | 2.64 | (1.65, 4.22) |
|  | First Treatment: NIPPV | 0.88 | 0.12 | 7.24 | 0.0000 | 2.42 | (1.9, 3.07) |
|  | Time x HFNO | -0.04 | 0.01 | -3.25 | 0.0011 | 0.96 | (0.94, 0.98) |
|  | Time x NIPPV | -0.02 | 0.01 | -2.27 | 0.0231 | 0.98 | (0.97, 1) |
|  | Age | -0.00 | 0.00 | -1.32 | 0.1883 | 1.00 | (0.99, 1) |
|  | BMI | 0.01 | 0.00 | 2.02 | 0.0435 | 1.01 | (1, 1.02) |
|  | Gender: Female | 0.24 | 0.08 | 3.03 | 0.0024 | 1.27 | (1.09, 1.49) |
|  | Ethnicity: Hispanic or Latino | -0.13 | 0.09 | -1.54 | 0.1239 | 0.87 | (0.74, 1.04) |
|  | Not White | -0.39 | 0.10 | -3.88 | 0.0001 | 0.68 | (0.56, 0.82) |
|  | Respiratory Rate (Breaths/Min) | -0.02 | 0.01 | -3.15 | 0.0016 | 0.98 | (0.97, 0.99) |
|  | SPO2/FIO2 | 0.01 | 0.00 | 8.05 | 0.0000 | 1.01 | (1, 1.01) |
|  | Transformed 1st Treatment Start (Hours After Hospital Admission) | 0.01 | 0.01 | 0.84 | 0.4025 | 1.01 | (0.99, 1.03) |
|  | Hospital Size: Medium | -0.68 | 0.20 | -3.37 | 0.0007 | 0.51 | (0.34, 0.75) |
|  | Hospital Size: Large | -0.91 | 0.20 | -4.54 | 0.0000 | 0.40 | (0.27, 0.6) |

**Figure 7:**
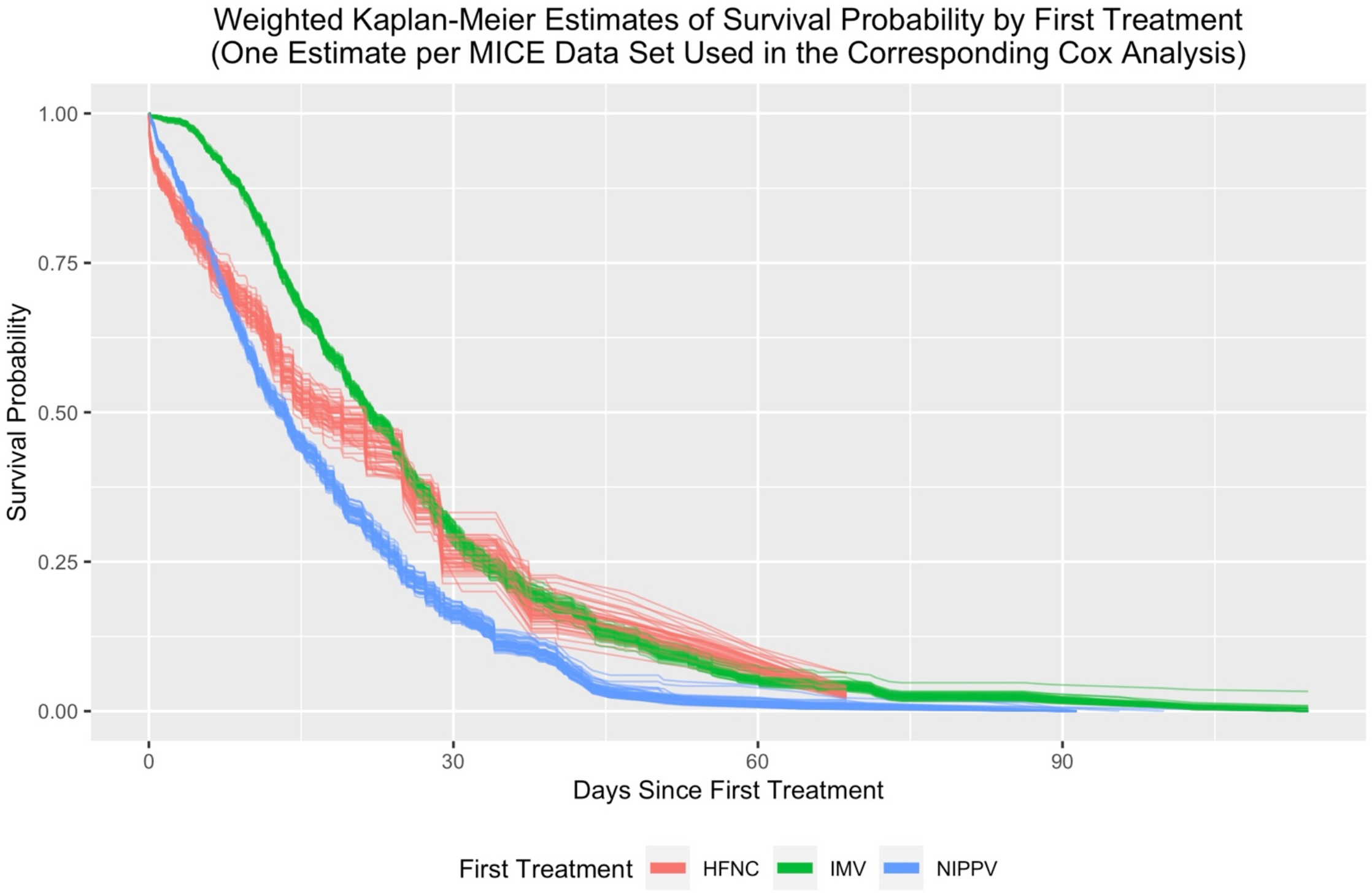
Time-to-Hospital Discharge Alive (HFNO vs NIPPV vs IMV). Time-to-hospital discharge alive weighted Kaplan-Meier curves for HFNO versus NIPPV versus invasive mechanical ventilation. Time zero is time of first treatment assignment. In these Kaplan-Meier estimates, people experiencing death are treated as censored. Curves use propensity scores estimated via the mean absolute standardized bias stopping rule in the generalized boosted models.

**Table 8:** Time-to-Hospital Discharge Alive (HFNO vs NIPPV vs IMV) Sensitivity Analysis. Death is treated as a competing risk. Time zero is time of hospital admission.

| Parameter | Estimate | SE | Wald Z Statistic | P-Value | Hazard Ratio | 95% CI on HR |
| --- | --- | --- | --- | --- | --- | --- |
| First Treatment: HFNO | 0.81 | 0.24 | 3.36 | 0.0008 | 2.25 | (1.4, 3.61) |
| First Treatment: NIPPV | 0.79 | 0.13 | 6.13 | 0.0000 | 2.21 | (1.71, 2.85) |
| Time x HFNO | -0.02 | 0.01 | -2.78 | 0.0055 | 0.98 | (0.96, 0.99) |
| Time x NIPPV | -0.01 | 0.01 | -1.72 | 0.0848 | 0.99 | (0.98, 1) |
| Age | -0.01 | 0.00 | -1.78 | 0.0747 | 0.99 | (0.99, 1) |
| BMI | 0.01 | 0.00 | 2.24 | 0.0253 | 1.01 | (1, 1.02) |
| Gender: Female | 0.22 | 0.08 | 2.68 | 0.0074 | 1.24 | (1.06, 1.46) |
| Ethnicity: Hispanic or Latino | -0.18 | 0.09 | -2.02 | 0.0432 | 0.84 | (0.71, 0.99) |
| Not White | -0.35 | 0.10 | -3.50 | 0.0005 | 0.71 | (0.58, 0.86) |
| Respiratory Rate (Breaths/Min) | -0.02 | 0.01 | -3.76 | 0.0002 | 0.98 | (0.97, 0.99) |
| SPO2/FIO2 | 0.01 | 0.00 | 8.95 | 0.0000 | 1.01 | (1, 1.01) |
| Transformed 1st Treatment Start (Hours After Hospital Admission) | -0.09 | 0.01 | -8.03 | 0.0000 | 0.92 | (0.9, 0.94) |
| Hospital Size: Medium | -0.60 | 0.21 | -2.88 | 0.0039 | 0.55 | (0.37, 0.83) |
| Hospital Size: Large | -0.78 | 0.21 | -3.79 | 0.0002 | 0.46 | (0.31, 0.69) |

**Figure 8:**
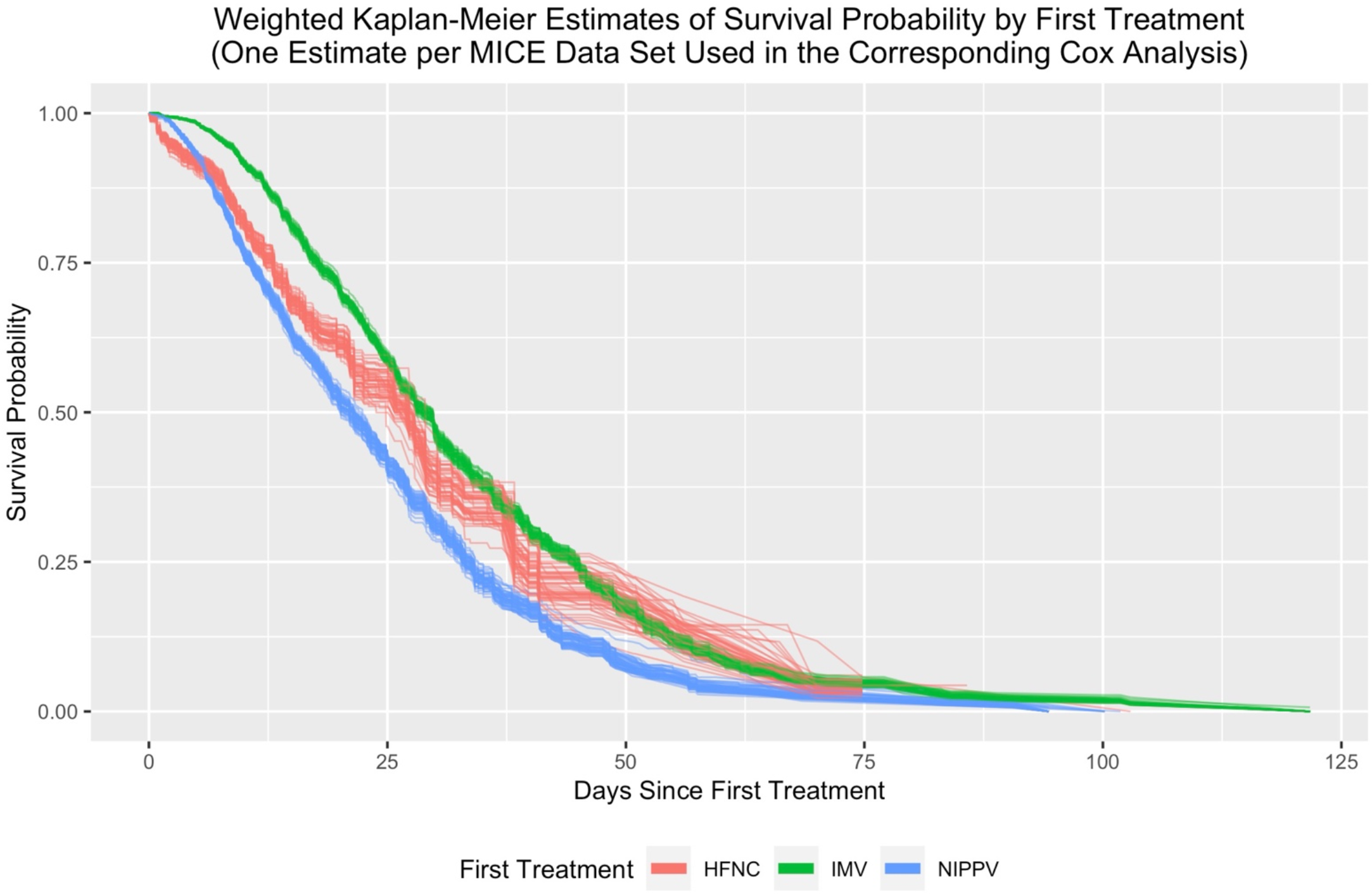
Time-to-Hospital Discharge Alive (HFNO vs NIPPV vs IMV) Sensitivity Analysis. Time-to-hospital discharge alive weighted Kaplan-Meier curves for HFNO versus NIPPV versus invasive mechanical ventilation. Time zero is time of hospital admission. In these Kaplan-Meier estimates, people experiencing death are treated as censored. Curves use propensity scores estimated via the mean absolute standardized bias stopping rule in the generalized boosted models.

**Table 9:**
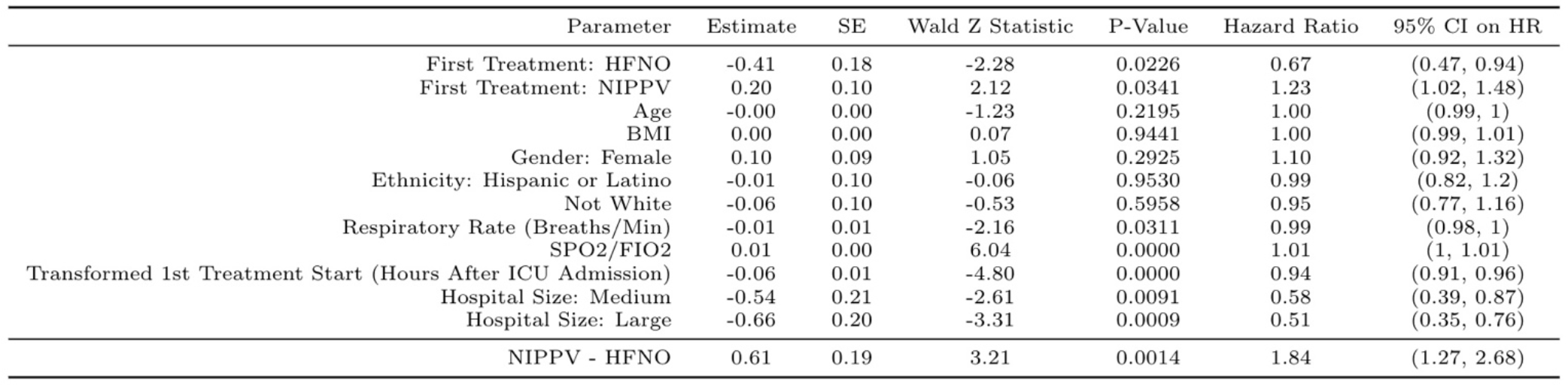
Time-to-ICU Discharge Alive (HFNO vs NIPPV vs IMV), with death as a competing risk for HFNO versus NIPPV versus invasive mechanical ventilation. Time zero is time of ICU admission. Model estimates shown below correspond to the mean absolute standardized bias stopping rule in the generalized boosted models. Inferential results were the same for the other stopping rules.

**Figure 9:**
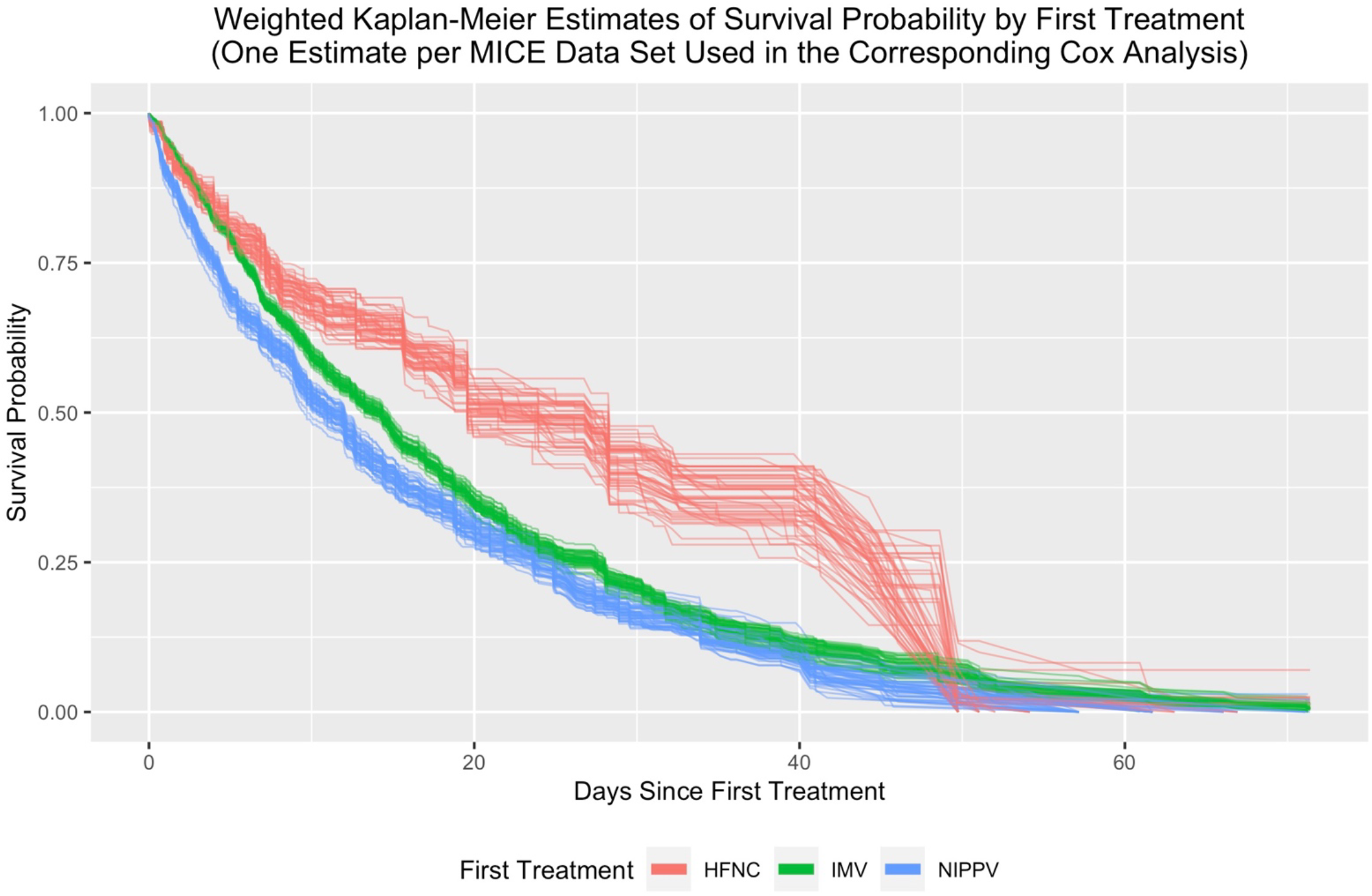
Time-to-ICU Discharge Alive (HFNO vs NIPPV vs IMV). Weighted Kaplan-Meier curves for HFNO versus NIPPV versus invasive mechanical ventilation. Time zero is time of ICU admission. In these Kaplan-Meier estimates, people experiencing death are treated as censored. Curves use propensity scores estimated via the mean absolute standardized bias stopping rule in the generalized boosted models.

STROBE Checklist

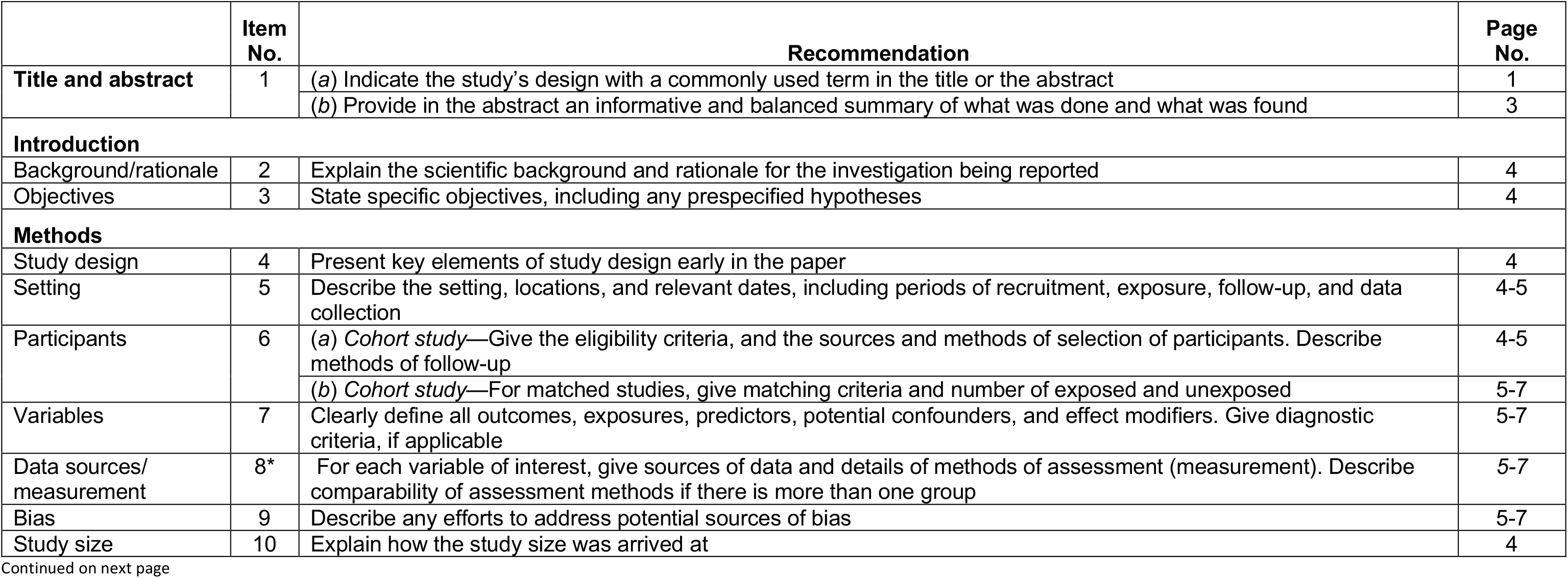
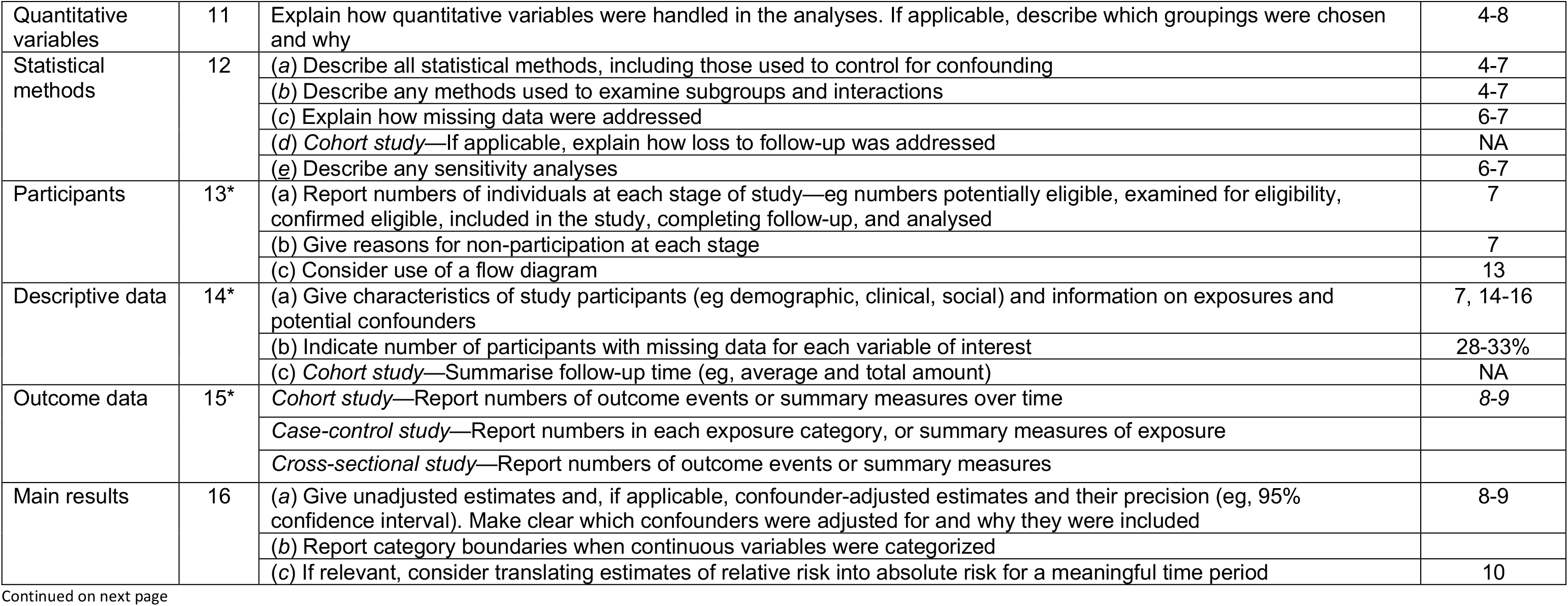
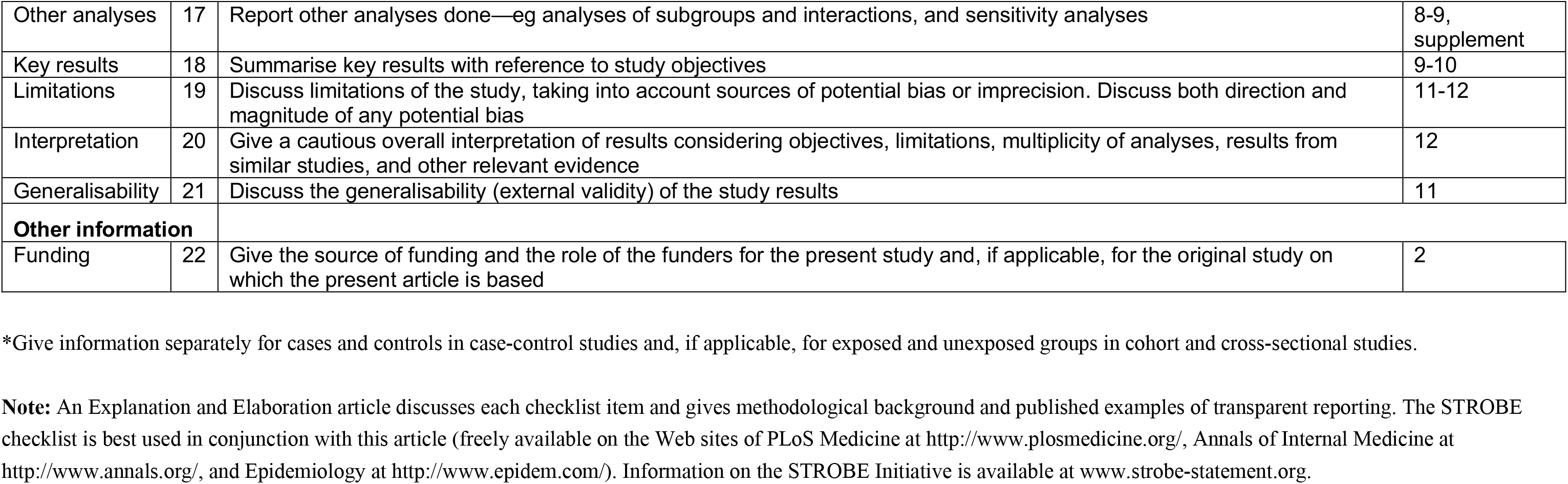

|  | Item No. | Recommendation | Page No. |
| --- | --- | --- | --- |
| Title and abstract | 1 | (a) Indicate the study's design with a commonly used term in the title or the abstract | 1 |
|  |  | (b) Provide in the abstract an informative and balanced summary of what was done and what was found | 3 |
| Introduction |  |  |  |
| Background/rationale | 2 | Explain the scientific background and rationale for the investigation being reported | 4 |
| Objectives | 3 | State specific objectives, including any prespecified hypotheses | 4 |
| Methods |  |  |  |
| Study design | 4 | Present key elements of study design early in the paper | 4 |
| Setting | 5 | Describe the setting, locations, and relevant dates, including periods of recruitment, exposure, follow-up, and data collection | 4-5 |
| Participants | 6 | (a) Cohort study—Give the eligibility criteria, and the sources and methods of selection of participants. Describe methods of follow-up | 4-5 |
|  |  | (b) Cohort study—For matched studies, give matching criteria and number of exposed and unexposed | 5-7 |
| Variables | 7 | Clearly define all outcomes, exposures, predictors, potential confounders, and effect modifiers. Give diagnostic criteria, if applicable | 5-7 |
| Data sources/measurement | 8* | For each variable of interest, give sources of data and details of methods of assessment (measurement). Describe comparability of assessment methods if there is more than one group | 5-7 |
| Bias | 9 | Describe any efforts to address potential sources of bias | 5-7 |
| Study size | 10 | Explain how the study size was arrived at | 4 |
| Quantitative variables | 11 | Explain how quantitative variables were handled in the analyses. If applicable, describe which groupings were chosen and why | 4-8 |
| Statistical methods | 12 | (a) Describe all statistical methods, including those used to control for confounding | 4-7 |
|  |  | (b) Describe any methods used to examine subgroups and interactions | 4-7 |
|  |  | (c) Explain how missing data were addressed | 6-7 |
|  |  | (d) *Cohort study*—If applicable, explain how loss to follow-up was addressed | NA |
|  |  | (e) Describe any sensitivity analyses | 6-7 |
| Participants | 13* | (a) Report numbers of individuals at each stage of study—eg numbers potentially eligible, examined for eligibility, confirmed eligible, included in the study, completing follow-up, and analysed | 7 |
|  |  | (b) Give reasons for non-participation at each stage | 7 |
|  |  | (c) Consider use of a flow diagram | 13 |
| Descriptive data | 14* | (a) Give characteristics of study participants (eg demographic, clinical, social) and information on exposures and potential confounders | 7, 14-16 |
|  |  | (b) Indicate number of participants with missing data for each variable of interest | 28-33% |
|  |  | (c) *Cohort study*—Summarise follow-up time (eg, average and total amount) | NA |
| Outcome data | 15* | *Cohort study*—Report numbers of outcome events or summary measures over time | 8-9 |
|  |  | *Case-control study*—Report numbers in each exposure category, or summary measures of exposure |  |
|  |  | *Cross-sectional study*—Report numbers of outcome events or summary measures |  |
| Main results | 16 | (a) Give unadjusted estimates and, if applicable, confounder-adjusted estimates and their precision (eg, 95% confidence interval). Make clear which confounders were adjusted for and why they were included | 8-9 |
|  |  | (b) Report category boundaries when continuous variables were categorized |  |
|  |  | (c) If relevant, consider translating estimates of relative risk into absolute risk for a meaningful time period | 10 |
| Other analyses | 17 | Report other analyses done—eg analyses of subgroups and interactions, and sensitivity analyses | 8-9, supplement |
| Key results | 18 | Summarise key results with reference to study objectives | 9-10 |
| Limitations | 19 | Discuss limitations of the study, taking into account sources of potential bias or imprecision. Discuss both direction and magnitude of any potential bias | 11-12 |
| Interpretation | 20 | Give a cautious overall interpretation of results considering objectives, limitations, multiplicity of analyses, results from similar studies, and other relevant evidence | 12 |
| Generalisability | 21 | Discuss the generalisability (external validity) of the study results | 11 |
| **Other information** |  |  |  |
| Funding | 22 | Give the source of funding and the role of the funders for the present study and, if applicable, for the original study on which the present article is based | 2 |

\*Give information separately for cases and controls in case-control studies and, if applicable, for exposed and unexposed groups in cohort and cross-sectional studies.
**Note:** An Explanation and Elaboration article discusses each checklist item and gives methodological background and published examples of transparent reporting. The STROBE checklist is best used in conjunction with this article (freely available on the Web sites of PLoS Medicine at http://www.plosmedicine.org/, Annals of Internal Medicine at http://www.annals.org/, and Epidemiology at http://www.epidem.com/). Information on the STROBE Initiative is available at www.strobe-statement.org.

